# Type 1 reaction leprosy patients display distinct immune-regulatory capacity before onset of symptoms

**DOI:** 10.1101/2023.12.18.23300119

**Authors:** Wilian Correa-Macedo, Monica Dallmann-Sauer, Marianna Orlova, Jeremy Manry, Vinicius M. Fava, Nguyen Thu Huong, Nguyen Ngoc Ba, Nguyen Van Thuc, Vu Hong Thai, Erwin Schurr

## Abstract

Leprosy is a chronic disease of the skin and peripheral nerves caused by *Mycobacterium leprae*. A major public health and clinical problem are leprosy reactions, which are inflammatory episodes that often contribute to nerve damage and disability. Type I reversal reactions (T1R) can occur after microbiological cure of leprosy and affect up to 50% of leprosy patients. Early intervention to prevent T1R and, hence, nerve damage, is a major focus of current leprosy control efforts. In a prospective study, we enrolled and collected samples from 32 leprosy patients before the onset of T1R. Whole blood aliquots were challenged with *M. leprae* sonicate or media and total RNA was extracted. After a three-year follow-up, the transcriptomic response was compared between cells from 22 patients who remained T1R-free and 10 patients who developed T1R during that period. Our analysis focused on differential transcript (i.e. isoform) expression and usage. Results showed that, at baseline, cells from T1R-destined and T1R-free subjects had no main difference in their transcripts expression and usage. However, the cells of T1R patients displayed a transcriptomic immune response to *M. leprae* antigens that was significantly different from the one of cells from leprosy patients who remained T1R-free. Transcripts with significantly higher upregulation in the T1R-destined group, compared to the cells from T1R-free patients, were enriched for pathways and GO terms involved in response to intracellular pathogens, apoptosis regulation and inflammatory processes. Similarly, transcript usage analysis pinpointed different transcript proportions in response to the *in-vitro* challenge of cells from T1R-destined patients. Hence, transcript usage in concert with transcript expression suggested a dysregulated inflammatory response including increased apoptosis regulation in the peripheral blood cells of T1R-destined patients before the onset of T1R symptoms. Combined, these results provided detailed insight into the pathogenesis of T1R.

**Author Summary:** The prevention and clinical management of type 1 reactions (T1R) remain an important unmet need to reduce nerve damage in leprosy patients. It is not known why 30-50% of leprosy patients will develop T1R. This knowledge gap underlies the need for a better mechanistic understanding of T1R that could lead to biomarker candidates to identify leprosy patients who are at high risk of developing T1R. Here, we used a prospective design in which leprosy patients were enrolled before the onset of T1R.Whole blood samples were obtained at enrollment, aliquots were left unstimulated or were stimulated *M. leprae* antigens and total RNA was extracted. Patients were followed for three years at which time 10 out of 32 participants had developed T1R. Subsequent transcript expression and usage analyses revealed that groups differed little in their isoform landscape at baseline. Following stimulation, transcriptomic response differences became pronounced. Transcripts with higher response in T1R group preferentially involved genes of intracellular defense and inflammatory pathways. Among these transcripts, non-coding ones had higher frequency in T1R. Our study provided new insights into the T1R pathogenesis by suggesting a role for non-coding transcripts into the immune dysregulations of T1R and providing additional candidate genes and their isoforms to be further investigated.

## Introduction

Leprosy, caused by *Mycobacterium leprae,* is a curable disease of the skin and peripheral nerves that leads to variable degrees of neuropathy [1]. Since the introduction of multidrug therapy towards the end of the last century, prevalence of leprosy has declined from approximately 7.5 million to approximately 200,000 cases globally, a number which is entirely accounted for by new cases detected each year [2]. Since the year 2000, the number of new cases has remained stable, which is unexpected since humans are the only known medically relevant hosts for *M. leprae* and cure upon treatment is close to 100%. In addition, it is possible that due to reduced leprosy detection efforts, the true number of new cases is higher than reported.

A major problem for clinical management of patients are leprosy reactions which are intense systemic inflammatory episodes. Type 1 reversal reactions (T1R) are the most frequent manifestations of these adverse host responses, afflicting approximately 30% to 50% of leprosy patients [3]. T1R may manifest themselves even after microbiological cure and are a main contributor to peripheral nerve damage and disability in leprosy [3, 4]. Despite encouraging progress in the definition of transcriptomic disease markers, there remains a lack of diagnostic tools to identify leprosy patients at risk of T1R and the reason why a subset of patients advances to T1R is unknown [5, 6]. The prevention of nerve damage, identification of T1R predisposing factors as well as the early recognition of patients at risk of T1R are major efforts of current leprosy control [7].

Employing a prospective study design, we investigated RNA transcript (i.e. isoform) expression levels and transcript usage following challenge with *M. leprae* sonicate with the aim of improving the mechanistic understanding of T1R. We also considered if changes at the isoform transcript level might reveal candidate druggable pathways for a more specific treatment to manage or shutdown T1R. The study of gene transcripts has the potential to reveal dynamics missed by gene-centric transcriptomic studies. Indeed, we found several transcripts that implicate genes previously not reported to be involved in T1R. On a mechanistic level, our study unambiguously showed that peripheral blood cells from those leprosy patients who will develop T1R suffer from an intrinsic dysregulated inflammatory response to *M. leprae* antigens. Given that T1R patients share key genetic risk factors with Parkinson’s disease and Crohn’s disease patients [8, 9], our observations may have implications beyond leprosy.

## Methods

### Ethics statement

The study received approval from the regulatory authorities of Ho Chi Minh City, Vietnam (So3813/UB-VX and 4933/UBND-VX) and the Research Ethics Board of the Research Institute at McGill University Health Centre in Montreal, Canada (REC98-041). Informed consent was obtained for all subjects participating in the study.

### Subjects

We studied 32 unrelated Kinh Vietnamese leprosy patients who were enrolled within 3 months of their leprosy diagnosis (S1 Fig). At the time of enrollment, all participants were T1R-free. The sample of patients was described previously by Orlova *et al.* [10]. We had obtained informed consent from all participants and had collected blood samples as described previously [10]. After enrollment, all patients had regular clinical check-ups for 3 years and during this time 10 patients had developed T1R. For the present follow-up investigation, we selected RNA samples with a RNA integrity number (RIN) > 7 [11]. All prospective T1R subjects were males with a minimum age > 15 years at leprosy diagnosis. We then selected RNA samples from leprosy patients that matched these criteria. Participants who developed T1R over the three-year follow-up period are referred to as T1R (N = 10) while leprosy patients who remained T1R-free are referred to as LEP (N = 22).

### Whole blood assay and RNA extraction

Details about biological samples, incubation time and materials were described previously [10, 12]. Briefly, at enrollment, blood samples were obtained and immediately processed. Whole blood was divided in two aliquots with one part being stimulated with *Mycobacterium leprae* sonicate (20 µg/ml) for approximately 32 hrs or incubated with medium only. Total RNA was extracted using LeukoLOCK leukocytes filters (Ambion) and RNeasy kit (Qiagen), quantified and quality assessed with Agilent 2100 Bioanalyzer (Agilent). All samples had RIN values > 8.5 when stored at -80 °C in 2008. Total RNA integrity was reassessed in 2016 and RNA samples with RIN > 7 were selected for library preparation.

### RNA sequencing and expression estimation

Poly-A RNA selection and RNA-Seq libraries were generated using the KAPA Stranded mRNA-Seq Kit (Roche, KK8421) and sequenced as 75-bp paired-end reads on Illumina HiSeq 4000 with an expected throughput of 50 million reads per library. Raw fastq files were quality-checked with FastQC v0.11.8 [13], RSeQC v2.6.1 [14] and processed with cutadapt v2.6 [15] to remove leftover adaptors and low-quality bases (≤ 20 Phred score) allowing for a minimum read size of 50bp. Sequences were aligned to the human reference genome (GRCh38.p13) with STAR v2.7.3a [16] and mapped reads projected to transcriptome coordinates (ENSEMBL release 102) [17]. BAM files were input to Salmon v1.4.0 [18] to obtain transcript expression estimates. Parameters used for STAR and Salmon are shown in the Supplementary methods (S1 File).

### RNA expression matrix and transcript filtering

For the differential transcript expression (DTE) analysis, text files containing estimated expression and transcript per million (TPM) values for 144428 annotated features, for each subject, were used to create two transcript level expression matrices in R v4.0.3 [19] using tximport v1.18.0 [20] and biomaRt v2.46.3 [21]. We adjusted Salmon’s estimated expression employing tximport’s methods “lengthScaledTPM” and “dtuScaledTPM” for the matrices used for differential transcript expression or differential transcript usage, respectively, as discussed by Love *et al.* [22]. We next ran low expression filtering for each matrix and removed transcripts with estimated expression < 200 read fragments or with Counts Per Million (CPM) < 4 in 70% of the libraries. For the transcript expression matrix exclusively, we also created a less stringent filtering (estimated expression < 10) to use with the function “calcNormFactors” (edgeR v3.32.1 [23]) to generate scaling normalization factors with the TMM method [24].

For differential transcript usage (DTU) analysis, beyond filtering by coverage depth, it was required that genes had at least two transcripts passing low expression filtering and that expression of each transcript represented a minimum of 10% of the total gene expression. The final number of testable transcripts was 25909, of which 23560 (corresponding to 11197 genes) were tested for differential transcript expression and 16413 (5765 genes) for differential transcript usage.

### Differential transcript expression and usage testing

The DTE matrix subset of 23560 transcripts and TMM scaling factors were input to limma’s voom v3.46.0 [25, 26] and the DTE analysis (Fig 1A) was carried out as published for gene-level analysis [27]. To account for multiple testing, we calculated adjusted *P*-values as implemented by stageR v1.12.0 [28]. Differentially expressed transcripts (DET) were defined as Log2 fold-change (Log_2_FC) ≥ 0.5 or Log_2_FC ≤ -0.5 and StageR FDR ≤ 5%.

**Fig 1.**
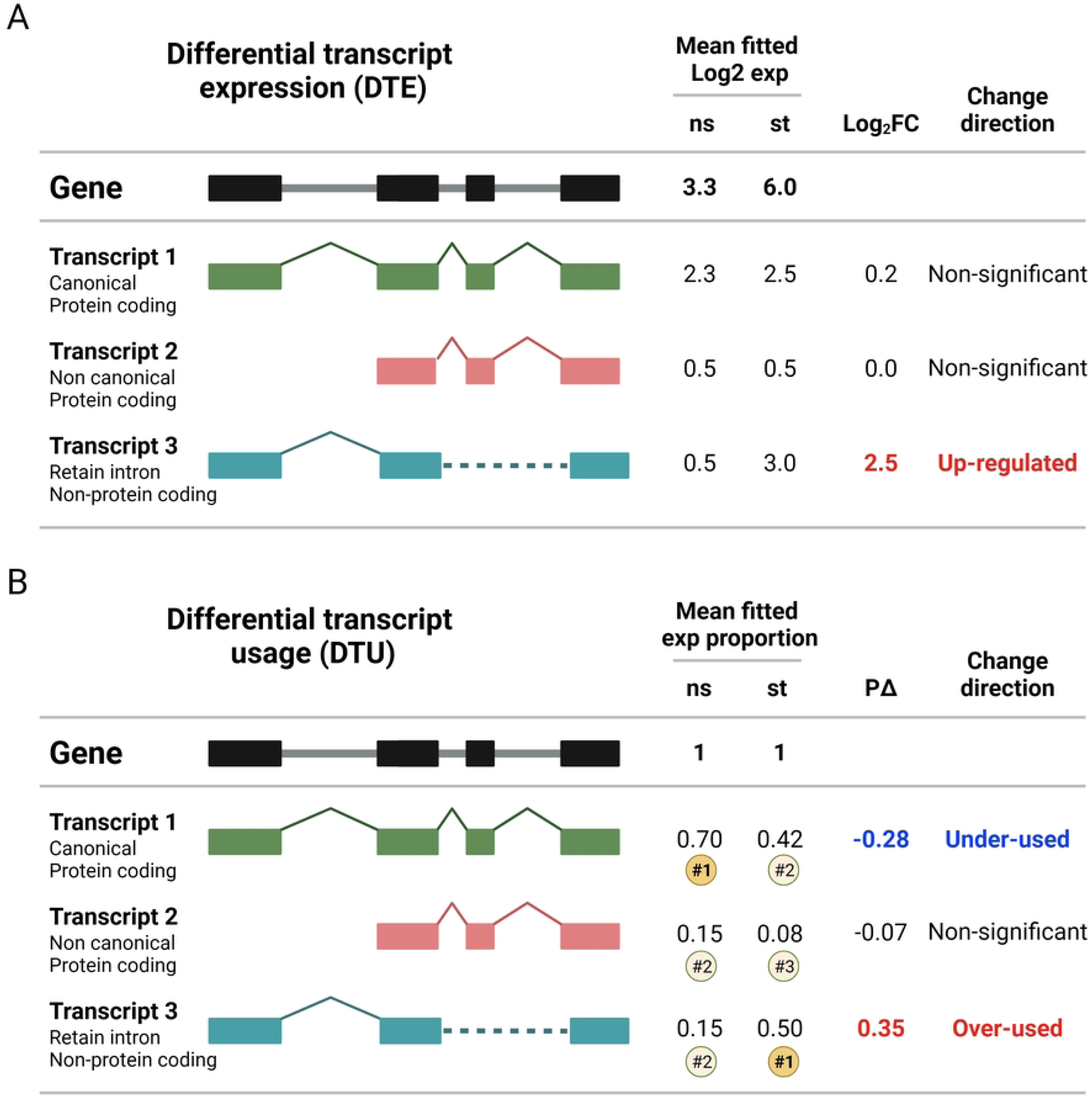
Schematic representations of differential transcript expression and differential transcript usage analyses. (**A-B**) On the left side of the figures, a schematic exon-intron structure of a gene is presented (black illustrations), and three possible transcripts resulting from alternative splicing are shown as colored illustrations. Boxes indicate exons while solid straight lines indicate intronic sequences that are expected to be spliced out. Chevrons indicate introns that must be spliced out to generate alternative mature RNAs and dashed lines indicate intron retention. The right half of the figures provides hypothetical examples for the two types of testing we employed. **(A)** Differential expression analysis was carried out for each transcript as independent genomic read-out. Expression differences between i) T1R vs LEP baseline (non-stimulated), ii) stimulated vs non-stimulated by group (e.g. LEP_st_ vs LEP_ns_) or iii) differential group responses ((T1R_st_ vs T1R_ns_) vs (LEP_st_ vs LEP_ns_)) were tested as Log_2_(CPM), yielding Log_2_FC as effect size for expression differences. In the example, libraries presented suggestive mean gene-level expression difference in response to stimulation. At transcript-level, however, only the non-protein coding (transcript 3) showed significant up-regulation. **(B)** For differential transcript usage analysis, normalized transcript expression was converted to proportion usage by dividing transcript expression by its parental gene expression. Differential transcript usage effect sizes are represented as the difference of mean fitted proportion usage (PΔ) between libraries. In the example from this figure, differential transcript usage shows that despite the overall expression increase after stimulation (panel A), the canonical transcript 1 had lower usage compared to baseline favoring the usage of the non-coding transcript. The numbers inside the circles show the ranking of the transcript proportions at baseline and in the stimulated samples. In this example, there is a switch of main transcripts between the two conditions, with transcript 1 ranked as the most common in the baseline and transcript 3 in the stimulated samples. Exp: expression; ns: non-stimulated samples; st: *M. leprae* sonicate stimulated samples.

DTU analysis (Fig 1B) was ran with DRIMSeq v1.18.0 [29] and executed as in a published workflow [22]. Unlike DTE analysis, in which Log_2_FC is used to compare the expression differences between groups, DRIMSeq does not provide an effect size for the usage analysis. To aid the identification of transcripts affected by stronger usage effects, we derived the mean fitted proportion usage for each transcript and library group and calculated the difference of means (referred as “proportion delta” or PΔ) for each pairwise contrast (Fig 1B). To determine an effect size cutoff, we pooled PΔ from all transcripts with StageR FDR ≤ 5% from all contrasts and calculated the median absolute deviation (MAD) which resulted in a rounded value of 0.07. Hence, transcripts were considered differentially used transcripts (DUT) when the effect size was PΔ ≥ 0.07 or PΔ ≤ -0.07 and StageR FDR ≤ 5%. We also determined transcript switching by considering the usage proportion of all testable transcripts under a gene. In such instances, transcripts may gain or lose their expression share “priority” relative to other transcripts from the same gene (Fig 1B). We only described transcript switching for DUT between the T1R and LEP groups responses to *M. leprae* sonicate stimulation (S2 Fig). Briefly, after DTU analysis, we ordered transcripts as a function of decreasing median proportion usage in a gene-wise fashion, for each library group (T1R_st_, T1R_ns_, LEP_st_, LEP_ns_). Next, we tested if the fitted proportion usage was statistically different among transcripts from a parental gene to assign ranking. Then we compared if the ranking of any transcript changed between library groups and confirmed that this change was also significant by retrieving the results from the DTU analysis. For transcripts that changed rank and were DUT, the corresponding genes were considered to contain a transcript usage switch event (S2 Fig).

### Regression models

Both DTE and DTU analyses relied on regression frameworks to detect differences between the T1R and LEP groups. Two models were used for DTE and one for DTU. The three models and their equations are presented in detail in the Supplementary methods (S1 File). Briefly, in the first model, we compared the baseline transcript expression between T1R and LEP, using all the libraries to estimate mean expression. To that end we declared stimulation status, age, RIN and sequencing batch as covariates, while a factor with group designation was the explanatory variable. In the second model, we analyzed the DTE in response to *M. leprae* stimulation (stimulated vs non-stimulated) for each group and then compared the response differences induced by stimulation between the LEP and T1R groups, via interaction analysis. To better account for the inter-individual variability, we used subject ID as fixed effect covariate and a nested term of group:stimulation, one per group, as explanatory variables (See Supplementary methods in S1 File). The third model, used for the DTU analyses, employed age, RIN and sequencing batch as covariates. A factor for group assignment, another for stimulation status and an interaction term of both were used as explanatory variables. For the baseline analysis of DTU, we compared the transcript usage of the non-stimulated samples between LEP and T1R. For the response to the stimulation, we compared the transcript usage between the stimulated vs non-stimulated samples by group and we compared the differential usage in the responses to stimulation between the two groups.

### Gene ontology and pathway enrichment analyses

Pathways and gene ontology (GO) enrichment analyses were performed with clusterProfiler package version 3.16.0 [30] and ReactomePA version 1.32.0 [31]. To carry out enrichment tests, we employed Ensembl or Entrez gene identifiers as proxies to tag biological functions of transcripts. Gene lists were derived from converting DET or DUT IDs into single gene IDs. Three different databases and functions were used for the enrichment analysis, KEGG was tested with enrichKEGG function, Reactome with enrichPathway and GO biological process was tested via enrichGO [32-35]. Three sets of analyses were run for each of the four contrasts investigated in DTE and DTU: i) any transcript change, irrespective of effect size direction; ii) positive Log_2_FC or PΔ values and iii) negative Log_2_FC or PΔ values. GO terms and pathways that had less than five assigned genes (proxies for significant transcripts) were excluded, and categories were considered significant if Benjamini-Hochberg FDR ≤ 10%.

## Results

### Differences in gene transcript expression and usage in whole blood from leprosy patients prior to onset of T1R

Employing a prospective study design, our sample consisted of 22 leprosy patients (LEP group) and 10 leprosy patients who developed T1R within three years of diagnosis (S1 Fig). A summary of the clinical and demographic data of the study population is shown in Table 1. T1R-destined and T1R-free leprosy patients were matched for age, which is a known T1R risk factor [36].

**Table 1.**
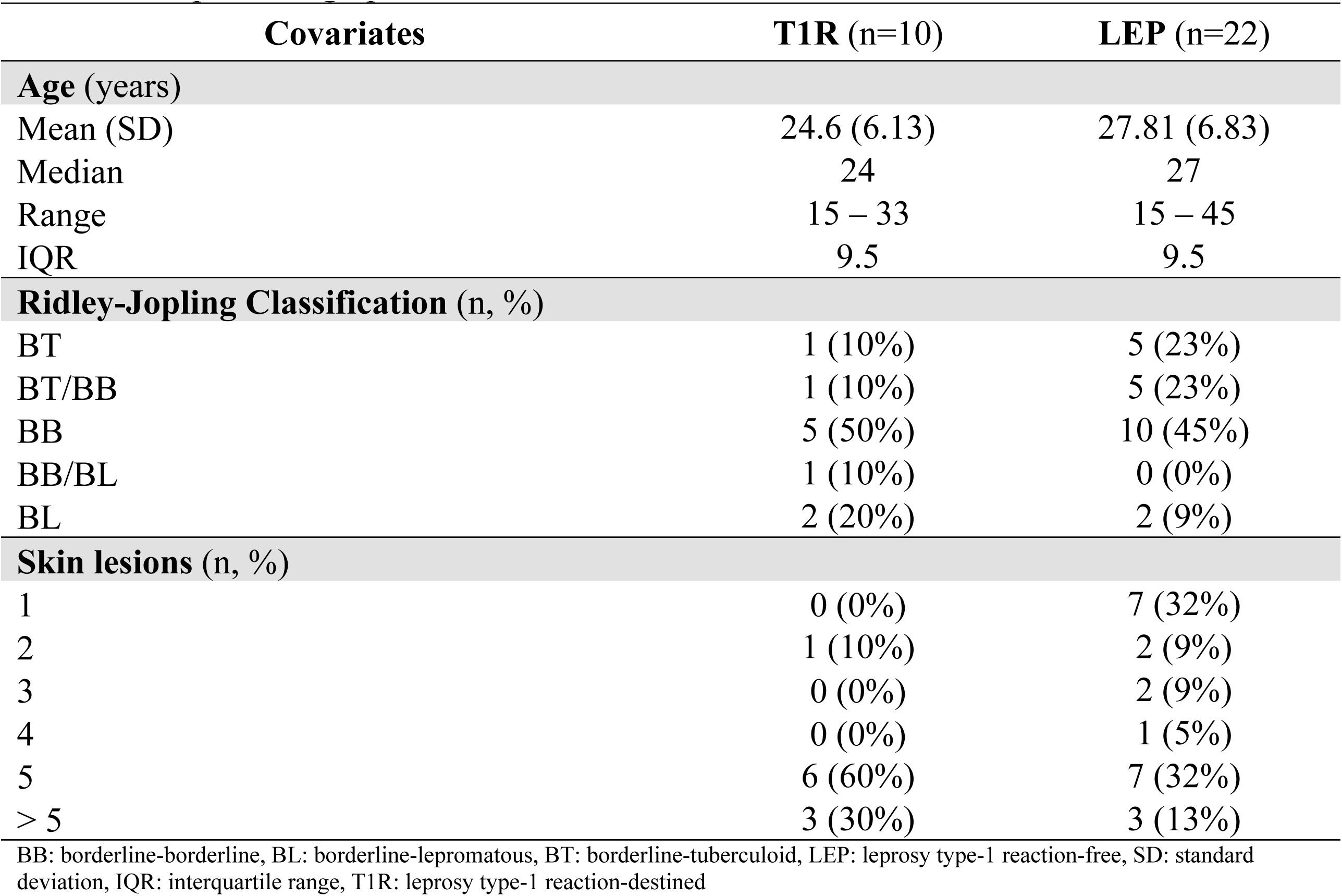
Sample demographics.

Enrolling participants prior to the onset of T1R symptoms allowed us to compare the transcriptomic response to *M. leprae* sonicate between the T1R and LEP groups in the absence of excessive inflammatory responses, characteristic of T1R episodes.

A total of 23560 transcripts were analyzed for DTE analysis and 16413 for DTU analysis at baseline and after *M. leprae* stimulation. For the DTE analysis, 25% (5909/23560) of tested transcripts were non-coding while for DTU this proportion was 30% (4922/16413). Comparing baseline transcript expression of the T1R against the LEP group resulted in 7 DET. However, these transcripts presented low average expression in both groups (S3A Fig). This suggested no main difference in the baseline transcript expression of blood cells from T1R and LEP. For baseline DTU analysis, T1R subjects differed from LEP subjects by 192 DUT that implicated 157 genes (S3B Fig and S1 Table). No GO term or pathway was significantly enriched with the parental genes from the DUT. Inspecting each of these transcripts for the annotated functions revealed that the majority encode proteins involved in general cellular processes such as intracellular trafficking, signal transduction and transcript processing. However, 20% (39/192) of the transcripts represented 33 genes that were linked to the immune response (e.g. *BIRC3*, *CMKLR1*, *COMMD6*, *NKIRAS1, NMT1, IFI16, PARP8*). Of these, 28 genes were implicated by coding transcripts (S1 Table). Two common themes for immune response genes tagged by DUT were apoptosis control and inflammatory responses. For example, *HIPK3* is a negative regulator of apoptosis [37] and we detected strong under-use of a protein coding *HIPK3* transcript in the T1R relative to the LEP group (PΔ = -0.20, S1 Table). Another example, *MS4A2*, encodes the beta subunit of the high affinity IgE receptor which is involved in allergic reactions [38]. Two DUT for *MS4A2* were detected with strong baseline usage differences between T1R and LEP participants, with T1R patients decreasing the usage of a protein coding transcript (PΔ = -0.35) in favor of increasing usage of a non-coding transcript (PΔ = 0.35, S1 table). Ascertaining whether these constitutive transcript usage differences for immune-related genes contribute to predisposition to T1R will require functional follow-up studies.

### Differential transcript expression in response to *in-vitro* challenge with *M. leprae* antigens

*M. leprae* antigen stimulation triggered similar numbers of DET for the LEP and T1R groups and their responses had similar profiles with up-regulation being the prominent effect (Fig 2A-B). Despite the response similarities, we also detected group-specific differences in the strength of the transcript response (Fig 2C-D). Overall, 6680 and 5932 transcripts were up- or down-regulated in response to *M. leprae* stimulus in at least one of the two groups (S2 Table), with similar number of DETs detected by group (Table 2). To identify biological processes that were affected by *M. leprae*-triggered transcript expression changes, for each group and response direction (i.e. positive Log_2_FC or negative Log_2_FC, Fig 2A-B), we performed GO term and pathway enrichment analyses using the parental genes from DETs (Table 2). When considering both up- and down-regulated transcripts in response to the *M. leprae* stimulus, a total of 1063 terms were observed for the LEP group and 837 terms for the T1R group. In both groups, term enrichment for the whole gene set was mainly driven by genes tagged by up-regulated transcripts (Table 2). Indeed, when focusing on genes tagged by up-regulated transcripts twice as many significant terms were detected (Table 2; S1 Data; S4 Fig). Most DET were shared between the LEP and T1R groups with the same direction of change by both groups for 84% (5639 / 6680) of up-regulated transcripts and for 67% (4014 / 5932) of down-regulated transcripts (Fig 2D). Testing GO terms or pathway enrichment with overlapping up-regulated transcripts yielded 1953 significant terms which revealed a prominent activation of immune response processes such as phagocytosis, antigen presentation, cytokine production and response, activation of innate immunity cell types, defense to intracellular pathogens and apoptosis (Fig 2E).

**Fig 2.**
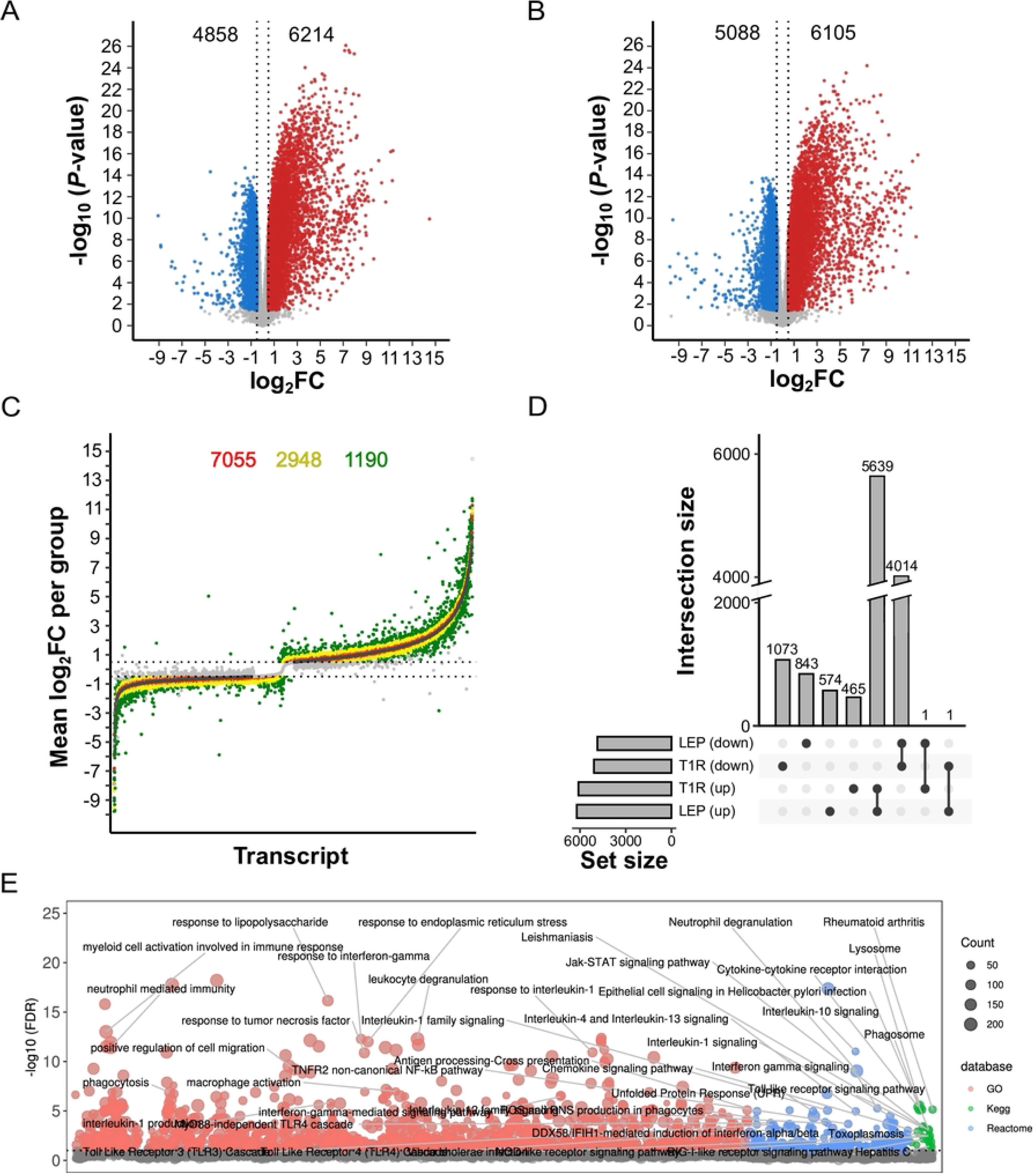
Transcript expression changes upon stimulation with *M. leprae* sonicate in whole-blood from leprosy T1R-free (LEP) and leprosy T1R-destined (T1R) individuals. (**A-B)** Volcano plots of transcriptional response to stimulation for (**A**) the LEP and (**B**) the T1R groups. Transcript responses are displayed as function of Log_2_ fold-change (Log_2_FC, x-axis) and negative log_10_ unadjusted *P*-value (y-axis). Each dot corresponds to a single transcript (i.e. isoform) and vertical dashed lines show the effect size threshold for absolute Log_2_FC ≥ 0.5. Differentially expressed transcripts are shown as red or blue dots that indicate up- or down-regulated transcripts significant at StageR FDR ≤ 5%, respectively. Grey dots are transcripts that did not reach significance (**C**) Strip plot presenting the effect size differences of significant transcript changes, expressed as Log_2_FC, presented in panels A and B. Transcripts are ordered along the x-axis according to their Log_2_FC for the LEP group. Colored dots indicate three levels of transcript response differences between LEP and the T1R group (|Log_2_FC _T1R_ - Log_2_FC _LEP_|) of ≤ 0.2 (red), 0.21 to 0.49 (yellow) and ≥ 0.5 (green). Their total numbers are shown on top. Light-gray dots represent non-significant transcripts and the horizontal lines mark the |Log_2_FC| ≥ 0.5 thresholds applied for group responses. (**D**) UpSet plot displaying the intersection of DET profiles in the *M. leprae* response by group. The bar chart at the bottom left indicates the total number of up-regulated or down-regulated transcripts for the two groups (four sets), while the bottom central panel identifies how sets intersect. The vertical upper bars indicate the corresponding intersection size (transcript counts) shown at the lower part of the graph. Connected dots indicate transcripts overlap between the T1R and LEP groups. Unconnected dots highlight group specific transcripts. (**E**) Manhattan plot for the results of pathway and GO term enrichment tests on 2919 genes implicated by 5639 up-regulated DET for both the T1R and LEP groups (panel D). The y-axis indicates the negative Log_10_ for BH adjusted *P*-values while the tested terms are arranged along the x-axis. The horizontal dashed line represents the 10% FDR cut-off for significant pathways/GO terms. Dots are sized as a function of the number of DET-derived genes in a term and colors represent the database from which the terms were obtained.

**Table 2.** Differential transcript expression and term enrichment tests summary.

| Test | Contrast | Direction | # of DETs | # of parental genes | # of GO terms/pathways |  |  |  |
| --- | --- | --- | --- | --- | --- | --- | --- | --- |
|  |  |  |  |  | Total | GO BP | Reactome | KEGG |
| Baseline | T1R vs LEP | Combined<sup>A</sup> | 7 | 7 | 0 | 0 | 0 | 0 |
|  |  | Higher expression | 7 | 7 | 0 | 0 | 0 | 0 |
|  |  | Lower expression | 0 | 0 | 0 | 0 | 0 | 0 |
| Per group stimulus effect | LEP.st vs LEP.ns | Combined<sup>A</sup> | 11072 | 6396<sup>B</sup> | 1063 | 895 | 78 | 90 |
|  |  | Up-regulated | 6214 | 3229 | 2093 | 1745 | 269 | 79 |
|  |  | Down-regulated | 4858 | 3375 | 7 | 0 | 7 | 0 |
|  | T1R.st vs T1R.ns | Combined<sup>A</sup> | 11193 | 6505<sup>B</sup> | 837 | 712 | 32 | 93 |
|  |  | Up-regulated | 6105 | 3159 | 2008 | 1679 | 256 | 73 |
|  |  | Down-regulated | 5088 | 3558 | 1 | 0 | 0 | 1 |
| Response differences between groups | T1R.st/ns vs LEP.st/ns | Combined<sup>A</sup> | 381 | 354<sup>B</sup> | 103 | 55 | 5 | 43 |
|  |  | Higher response<sup>D</sup> | 207 | 198 | 81 | 26 | 13 | 42 |
|  |  | Lower response<sup>D</sup> | 174 | 162 | 8 | 8 | 0 | 0 |

<sup>A</sup> Combined count is the sum of the two directions of Log<sub>2</sub>FC (i.e. up-regulated and down-regulated).
<sup>B</sup> Each gene implicated by more than one DET present in the two directions is counted once. Hence, the “Combined” count of genes does not equal the sum of genes by direction.
<sup>C</sup> Count of transcripts used for gene ontology analyses. Only terms with FDR ≤ 10% and with at least 5 DET-derived genes assigned to them were considered significant.
<sup>D</sup> Higher and lower responses refer to the positive (Log<sub>2</sub>FC<sub>T1R</sub> > Log<sub>2</sub>FC<sub>LEP</sub>) and negative (Log<sub>2</sub>FC<sub>T1R</sub> < Log<sub>2</sub>FC<sub>LEP</sub>) Log<sub>2</sub>FC from the interaction test, respectively.
DET: Differentially expressed transcripts; GO BP: Gene ontology biological processes; LEP: leprosy group (T1R-free); ns: non-stimulated; st: stimulated; T1R: Type-1 reaction group (T1R-destined).

Next, we checked for group response differences by comparing the effect sizes for transcripts that were differentially expressed in at least one group. We found that the bulk of DET presented similar effect sizes for both groups (shown in red and yellow in Fig 2C). DET with absolute Log_2_FC < 0.2 response difference between groups (red dots in Fig 2C), despite a large number of nominally significant transcripts, were significantly enriched in only a single “Ion channel transport” GO term. The 2948 DET with intermediate effect size difference (|Log_2_FC| ≥ 0.2 and < 0.5, yellow dots in Fig 2C) were enriched in 735 terms linked to general inflammatory and intracellular defense to pathogens (S5A Fig). These terms were skewed towards neutrophil/myeloid cell activation and cell migration. Finally, 1190 out of 12612 transcripts presented effect size differences larger than |Log_2_FC| ≥ 0.5 (green dots, Fig 2C). These transcripts with stronger effect in one of the groups were evenly split between the T1R and LEP groups for up-regulated transcripts (Fig 2C, upper half). However, transcripts with stronger down-regulation (64%; 3798 / 5932; Fig 2C, lower half) were more frequent among T1R subjects. The whole 1190 DET set, resulted in 954 significant terms linked to innate immunity, cytokine production and cell motility (S5B Fig).

Of the 2955 transcripts that were differentially expressed in only one of the two groups, 465 DET were exclusively up-regulated for T1R resulting in 201 significant terms, while the term number for the 574 exclusive DET for LEP was 291. Despite being derived from DET exclusive lists for each group, 107 GO/pathway terms overlapped (S6 Fig). Even for group-specific up-regulated DET, the majority of terms that were specific for each group related back to the overlapping immunological processes (e.g. apoptosis). As for group-specific down-regulated DET, no GO/pathway terms were significant.

The bulk of DET response differences between T1R and LEP patients induced by *M. leprae* antigens relied, however, on transcripts that were shared between groups and up-regulated (n = 5639, representing 1953 significant terms, Fig 2D-E). For those, we observed that DETs had larger effects sizes for T1R in many terms related to immune response, while terms related to myeloid/neutrophil activation had higher response for LEP (S7 Fig). For overlapping down-regulated transcripts (n = 4014, Fig 2D), T1R also presented larger effects (stronger down-regulation). Despite the large number of genes tagged by down-regulate transcripts, the number of significant terms was drastically lower at 11. These terms revealed that down-regulated transcripts were involved in the cross talk between lymphoid and non-lymphoid cells, CD8 cytotoxicity, leukocyte transendothelial migration and chromatin organization.

Next, we employed a more stringent approach and formally tested the significance of the differential *M. leprae* transcriptomic responses between cells from T1R and LEP subjects (interaction DET). We detected 381 DET, implicating 354 genes, which responded after stimulation for at least one group and had a significant response difference of |Log_2_FC| ≥ 0.5 (Table 2, S2 Table, Fig 3A-B). These DET were distributed in six categories based on the direction of their individual group Log_2_FC (S8A Fig). The category with most interaction DET was up-regulation for both groups, with the T1R group presenting higher up-regulation (Figs 3C and S8B-F). Encompassing 38% (145/381) of interaction DET, differences captured in this category were enriched in terms related to intracellular defense, apoptotic signaling, and inflammatory response (Fig 3D). For instance, genes underlying the enrichment of inflammatory response terms included potent pro-inflammatory genes such as *IL12B* and *IL6* as well as genes with modulatory effects like *TNIP1* and *IL1RN*. These are example of genes that are known regulators of interleukin-1, interferon (IFN)-γ and NF-κB signaling pathways (Fig 3E). *IL12B* presented a single DET (Log_2_FC_T1R_ = 10.3; Log_2_FC_LEP_ = 8.1). *IL6* presented four DET, all more strongly up-regulated by T1R, however only the protein coding ENST00000258743 passed the test for significance (Log_2_FC_T1R_ = 9.8; Log_2_FC_LEP_ = 7.8, S2 and S3 tables). *TNIP1* had five up-regulated transcripts, of which two had significant group response differences, ENST00000315050 (Log_2_FC_T1R_ = 6.6; Log_2_FC_LEP_ = 4.3) and the non-coding ENST00000522574 (Log_2_FC_T1R_ = 2.8; Log_2_FC_LEP_ = 2.2). For *IL1RN*, only the non-coding ENST00000472292 was significantly different between T1R and LEP.

**Fig 3.**
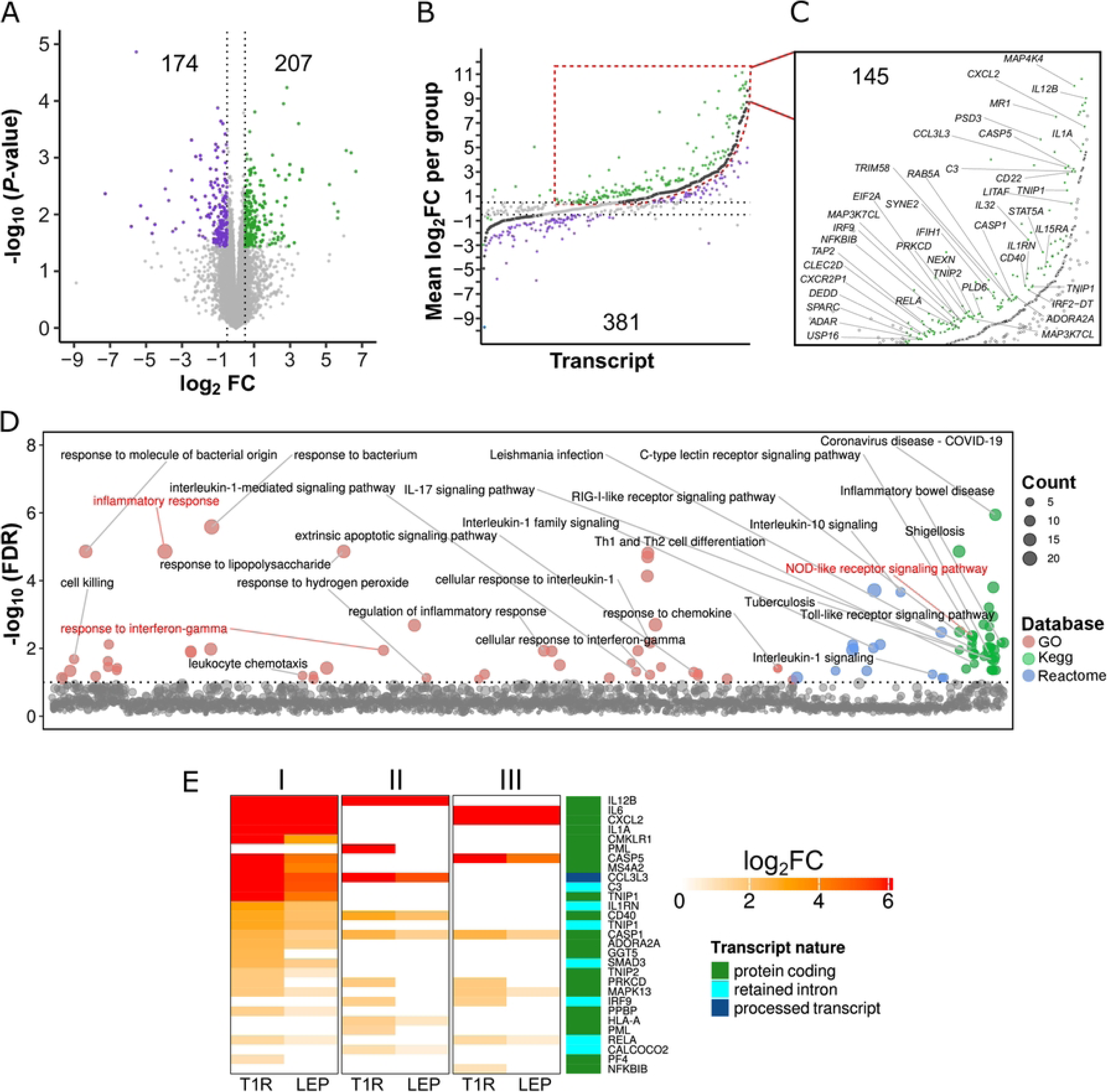
Following stimulation with *M. leprae* sonicate, cells from T1R-destined leprosy patients show increased transcriptional inflammatory responses relative to T1R-free leprosy patients. (**A**) Volcano plot of significant differential transcriptional response to *M. leprae* sonicate between the T1R and LEP groups detected via interaction analysis. Contrasting the Log_2_FC from T1R against LEP, for 23,560 tested transcripts, we identified 381 transcripts with significant differential response between the two groups. Light gray dots represent transcripts with non-significant response differences between groups, purple dots indicate significant responses with negative value for the contrast (Log_2_FC_T1R_ - Log_2_FC_LEP_), while green dots indicate a significant positive value. **(B)** Strip plot categorizing significantly different responses to *M. leprae* of the T1R and LEP groups for the 381 significant transcripts from panel **A**. Dark grey dots at the center represent 381 transcripts for the LEP group response sorted by increasing Log_2_FC_LEP_ values. They serve as reference for the significantly different response by T1R. Green or purple dots represent the Log_2_FC_T1R_ responses that underlie the positive or negative Log_2_FC for the contrast Log_2_FC_T1R_ - Log_2_FC_LEP_ observed in panel A, respectively. **(C)** Break-out focus of transcripts that were up-regulated by both the T1R and LEP groups with stronger up-regulation by T1R. Labeled dots are examples for increased T1R response by transcripts belonging to genes with immune/inflammatory functions. **(D)** Manhattan plot for the results of pathway and GO term enrichment tests on genes implicated by the 145 DET from panel C. The y-axis indicates the negative Log_10_ for BH-adjusted *P*-values while the tested terms are arranged along the x-axis. The horizontal dashed line represents the 10% FDR cut-off for significant pathways/GO terms. Dots are sized as a function of the gene number in a term and colors represent the database from which the terms were obtained. Immune response and inflammation terms are identified. Terms highlighted in red were selected to be further characterized in panel E. (**E**) Heatmap for 29 transcripts with significant differential transcript upregulation in response to *M. leprae* sonicate between T1R and LEP patients. The genes are enriched in the terms I) inflammatory response, II) response to IFN-γ, and III) NOD-like receptor signaling pathway. Rows represent the same transcript across the three main terms. Each term column is composed of two sub-columns representing the stimulation responses for the T1R or LEP groups. Colors represent the response intensity of transcripts as captured by Log_2_FC_T1R_ and Log_2_FC_LEP_ values, with stronger responses being depicted by scales of red. Transcripts were arranged along the y-axis by increasing Log_2_FC observed for the T1R group. For each row, transcripts that were not DET for a group or are not part of the term were assigned zero as Log_2_FC. The right most column identifies the transcript class.

Interestingly, out of the 145 DET with higher up-regulation in T1R (Fig 3C), 49 were non-coding transcripts, of which six represented genes encoding proteins with immune functions (*C3, CCL3L3, CLEC2D, IL1RN, IRF9* and *RELA*) (S9 Fig, and section A in S3 Table). *IRF9* (ENSG00000213928), for instance, presented two coding and four non-coding up-regulated transcripts in response to *M. leprae* sonicate with higher effect size for T1R (S2 Table). Yet, only the non-coding transcript ENST00000560365 showed a significant Log_2_FC = 0.98 response difference (S3 Table). Hence, we tested to what extent non-coding transcripts were contributing to the GO term/pathway enrichment results in Fig 3D. After removing all gene IDs implicated exclusively by non-coding DET, thereby reducing the input list from 137 to 92 genes, we obtained an increased number of immune-related terms whilst increasing the statistical significance for previously detected terms (S4 Table).

On the other hand, a total of 75 DET were more strongly up-regulated by LEP patients (S8F Fig and S3 Table). Interestingly, 14 out of these 75 interaction DET were responsible for over-representation of terms related to myeloid/neutrophil activation processes (S3 Table). Generally, the cells from the LEP group displayed a stronger transcriptional response of genes encoding modulators of inflammatory and apoptotic processes (e.g. *LAIR1*, *DOK3*, *MAGT1*, *FOXP1, OLFM4*, *OLR1*, *PRKCD, IL18BP*, and *NRBF2*) as well as pathogen recognition receptors (*CFP*, *CD14* and *TLR5*; S3 Table). Noteworthy, *PRKCD,* a gene coding for a pro-apoptotic factor, displayed two interaction DET (ENST00000330452 and ENST00000654719) both translating into proteins with identical sequence. The first is canonical transcript with longer 5’ and 3’ UTRs that was up-regulated and DET only for LEP, while the second with shorter UTRs was exclusively up-regulated for T1R (S3 Table). The remaining four categories from Fig 3B did not show enrichment in biological terms (S8B-E Fig).

Finally, we investigated whether the six main transcript response categories (S8A Fig) presented any imbalance of transcript type (Table 3). Among the down-regulated transcripts, the T1R group showed increased numbers of down-regulated canonical transcripts, although the frequency distribution was non-significant by Fisher’s test (Table 3). However, DET with higher up-regulation in the T1R group presented significantly increased proportion of non-coding vs coding DET (*P* = 0.0065, Table 3). The increased proportion of up-regulated non-coding transcripts among T1R subjects was primarily driven by transcripts annotated as retained intron and lncRNA (*P* = 0.0062 and 0.0326, Table 3). This analysis showed that, in terms of frequency, canonical transcripts were the least responsive transcript type differently changing between T1R and LEP in our experiments. Considering the regulatory role of non-coding RNA categories [39, 40], the significant frequency excess of these DET in the response mounted by T1R may play a role in the immune dysregulation in this group.

**Table 3.**
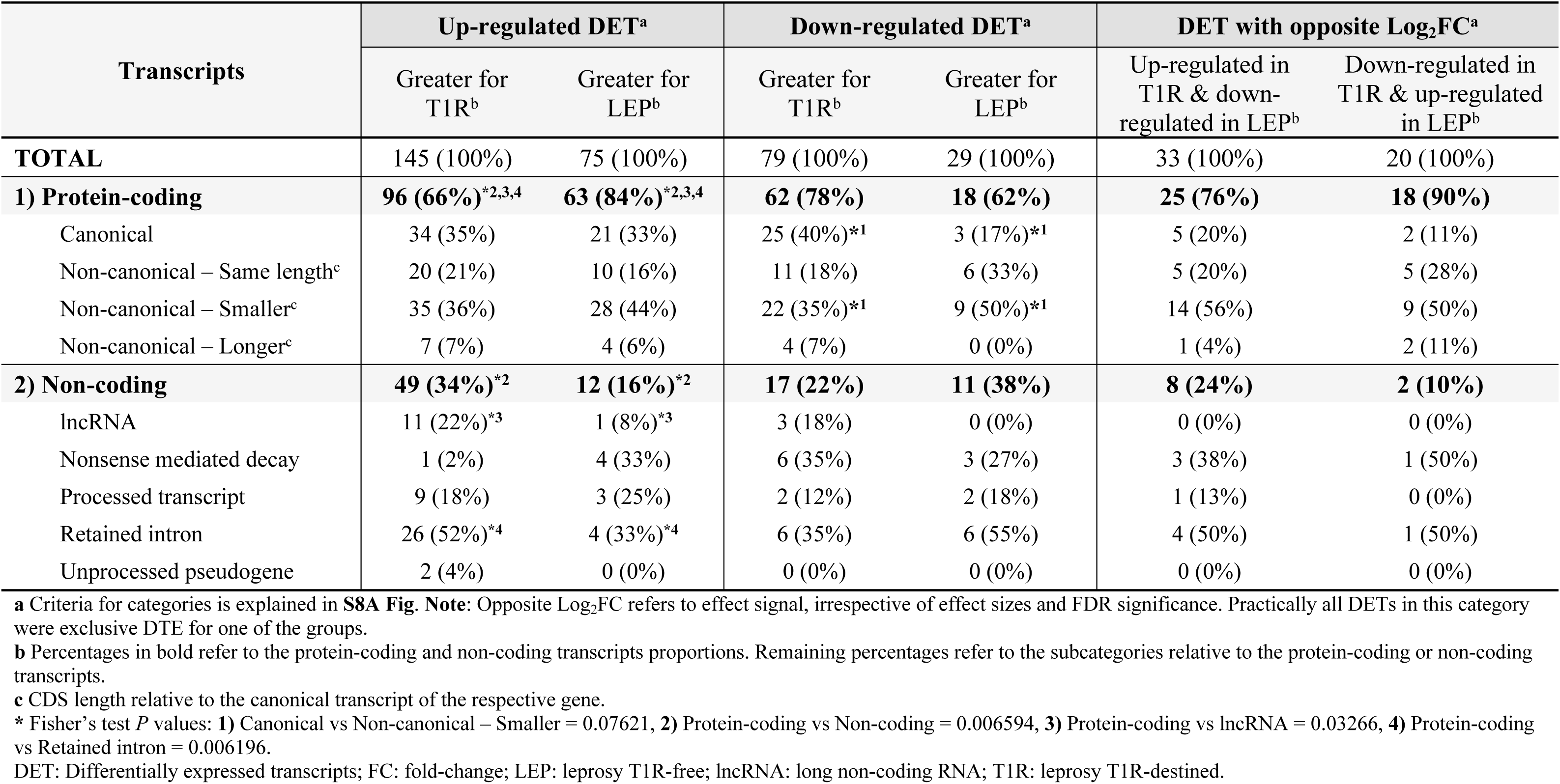
Distribution of transcripts by coding nature and coding sequence (CDS) length for interaction DET.

| Transcripts | Up-regulated DET<sup>a</sup> |  | Down-regulated DET<sup>a</sup> |  | DET with opposite Log<sub>2</sub>FC<sup>a</sup> |  |
| --- | --- | --- | --- | --- | --- | --- |
|  | Greater for T1R<sup>b</sup> | Greater for LEP<sup>b</sup> | Greater for T1R<sup>b</sup> | Greater for LEP<sup>b</sup> | Up-regulated in T1R & down-regulated in LEP<sup>b</sup> | Down-regulated in T1R & up-regulated in LEP<sup>b</sup> |
| **TOTAL** | 145 (100%) | 75 (100%) | 79 (100%) | 29 (100%) | 33 (100%) | 20 (100%) |
| **1) Protein-coding** | **96 (66%)\*<sup>2,3,4</sup>** | **63 (84%)\*<sup>2,3,4</sup>** | **62 (78%)** | **18 (62%)** | **25 (76%)** | **18 (90%)** |
| Canonical | 34 (35%) | 21 (33%) | 25 (40%)*<sup>1</sup> | 3 (17%)*<sup>1</sup> | 5 (20%) | 2 (11%) |
| Non-canonical – Same length<sup>c</sup> | 20 (21%) | 10 (16%) | 11 (18%) | 6 (33%) | 5 (20%) | 5 (28%) |
| Non-canonical – Smaller<sup>c</sup> | 35 (36%) | 28 (44%) | 22 (35%)*<sup>1</sup> | 9 (50%)*<sup>1</sup> | 14 (56%) | 9 (50%) |
| Non-canonical – Longer<sup>c</sup> | 7 (7%) | 4 (6%) | 4 (7%) | 0 (0%) | 1 (4%) | 2 (11%) |
| **2) Non-coding** | **49 (34%)\*<sup>2</sup>** | **12 (16%)\*<sup>2</sup>** | **17 (22%)** | **11 (38%)** | **8 (24%)** | **2 (10%)** |
| lncRNA | 11 (22%)*<sup>3</sup> | 1 (8%)*<sup>3</sup> | 3 (18%) | 0 (0%) | 0 (0%) | 0 (0%) |
| Nonsense mediated decay | 1 (2%) | 4 (33%) | 6 (35%) | 3 (27%) | 3 (38%) | 1 (50%) |
| Processed transcript | 9 (18%) | 3 (25%) | 2 (12%) | 2 (18%) | 1 (13%) | 0 (0%) |
| Retained intron | 26 (52%)*<sup>4</sup> | 4 (33%)*<sup>4</sup> | 6 (35%) | 6 (55%) | 4 (50%) | 1 (50%) |
| Unprocessed pseudogene | 2 (4%) | 0 (0%) | 0 (0%) | 0 (0%) | 0 (0%) | 0 (0%) |

**a** Criteria for categories is explained in **S8A Fig. Note:** Opposite Log<sub>2</sub>FC refers to effect signal, irrespective of effect sizes and FDR significance. Practically all DETs in this category were exclusive DTE for one of the groups.
**b** Percentages in bold refer to the protein-coding and non-coding transcripts proportions. Remaining percentages refer to the subcategories relative to the protein-coding or non-coding transcripts.
**c** CDS length relative to the canonical transcript of the respective gene.
\* Fisher's test *P* values: **1)** Canonical vs Non-canonical – Smaller = 0.07621, **2)** Protein-coding vs Non-coding = 0.006594, **3)** Protein-coding vs lncRNA = 0.03266, **4)** Protein-coding vs Retained intron = 0.006196.
DET: Differentially expressed transcripts; FC: fold-change; LEP: leprosy T1R-free; lncRNA: long non-coding RNA; T1R: leprosy T1R-destined.

### Differential transcript usage in response to *M. leprae* antigens

In the preliminary analysis, the numbers of DUT following *M. leprae* sonicate stimulation were similar between the cells from the T1R and LEP groups (S10 Fig). Overall, a total of 3568/15413 (21%) transcripts altered usage after stimulation (S10 Fig and S5 Table). From these, most transcripts (2836/3568, 79%) presented similar usage changes in response to the challenge between the two groups (red dots in S10 Fig). For all DUT, performing GO term and pathway enrichment analyses did not yield enrichment in any term. However, 36% (1281/3568) of the DUT parental genes were involved in immune/inflammatory responses such as TNF, NF-κB and multiple interleukins (pro and anti-inflammatory) signaling, recognition of intracellular pathogens and apoptosis. S11 Fig depicts the mean DUT response for 150 transcripts with the highest effect size difference between groups that are linked to GO terms for inflammatory response. This figure shows how sparse DUT profiles were for the broad GO terms “inflammatory response”, “innate immune response” and “apoptotic process”, which presented 45, 27 and 114 DUT, respectively. The profiles suggested that stimulated cells from both groups were setting up inflammatory and apoptosis processes, but transcripts implicating *TNFAIP3*, *NLRP2*, *PIDD1*, *HIPK3*, *GRAMD4*, *RSL1D1*, *TFDP2* and *CD40* (S11 Fig) suggested it would unfold via different routes in the two groups.

Formal testing for significant differential usage between T1R and LEP groups yielded 326 DUT (Fig 4A and S5 Table). Of these, we found 177 DUT (159 genes) with a transcript usage switch in at least one of the groups (S5 Table). DUT with switches in both groups represented the minority of cases (14 out of 177) while 59% (105/177) of usage switches occurred only in the T1R group and 33% (58/177) only in the LEP group. Despite the lack of significant GO term or pathway enrichment for any DUT subset, many implicated genes had important immune or regulatory functions. A graph presenting significant response differences in transcript usage by groups, their implicated genes, and tagged pathways is presented in S12 Fig.

**Fig 4.**
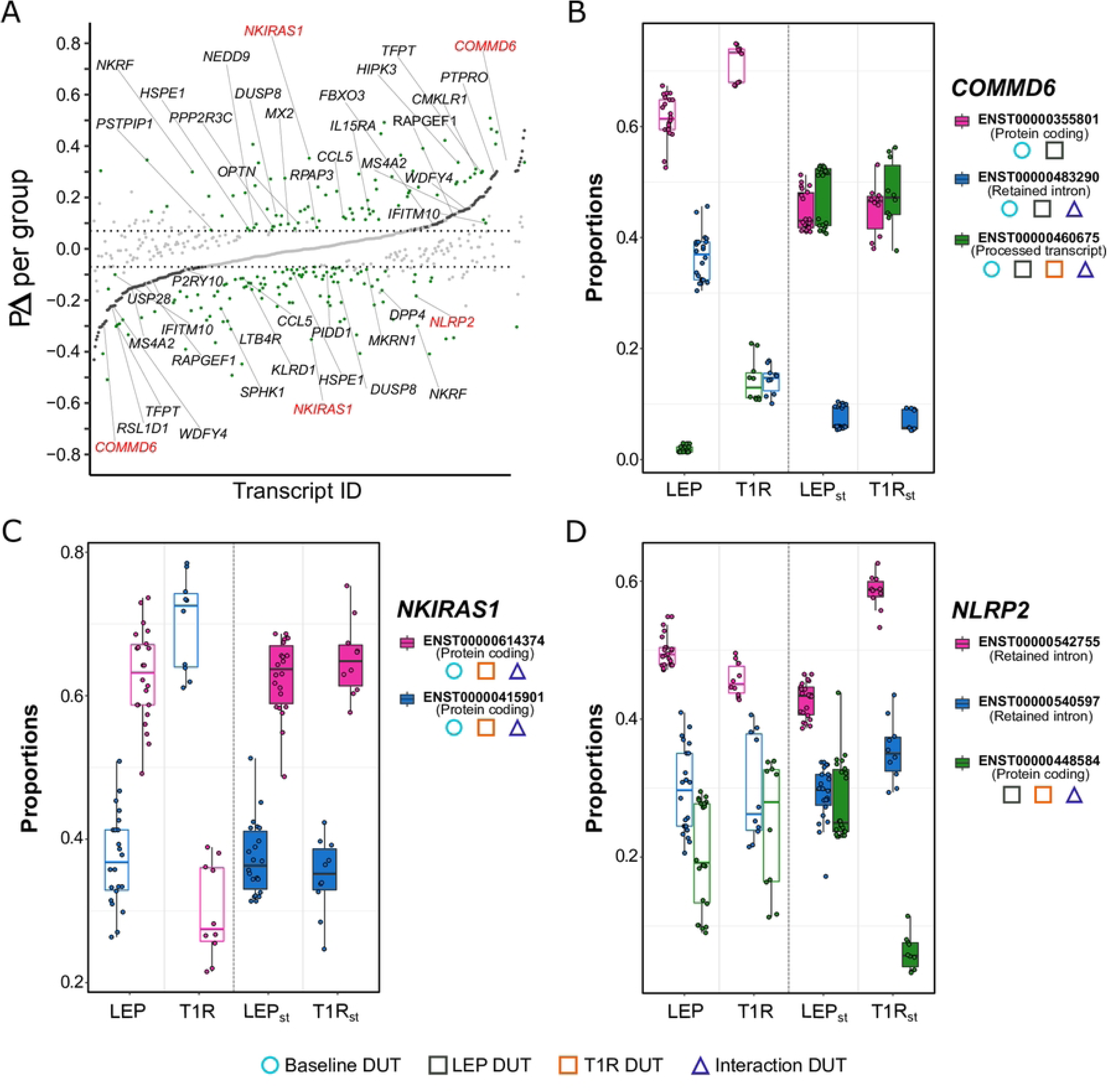
Significant differences in transcript usage in response to *M. leprae* sonicate between the cells from the T1R and LEP groups. **(A)** Strip plot for 326 transcripts with significant usage differences for |PΔ _T1R_ – PΔ _LEP_| ≥ 0.07. This plot presents the distribution of the mean fitted proportion difference (PΔ) in transcript usage by contrasting *M. leprae* sonicate stimulated against non-stimulated samples, for each group, and highlights the extent of differences between groups. Transcripts are represented by dots and ordered along the x-axis according to increasing PΔ values for the LEP group. Light gray dots represent transcripts with non-significant PΔ for both groups and horizontal lines mark the | PΔ | ≥ 0.07 thresholds. Differentially used transcripts (DUT) for LEP are depicted as dark-grey dots and appear as a continuous curve in the center part of the graph while green dots display the PΔ for the T1R group response. Transcripts identified by black gene symbols are involved in immune/inflammatory/apoptotic processes, while red symbols display examples selected to show details of transcript usage in panels B to D. **(B-D)** Boxplots presenting the fitted proportion usage for transcripts from three genes that encode regulators of the NF-κB signaling pathway: (**B**) *COMMD6*, (**C**) *NKIRAS1* and (**D**) *NLRP2*. Fitted proportions are shown as dots representing values for each subject. Open boxes represent distribution for non-stimulated samples, while closed boxes refer to *M. leprae* sonicate stimulation (LEP_st_ and T1R_st_). Transcript IDs are displayed to the right of each plot and symbols shown below the transcript IDs identify transcripts that were DUT for the baseline (light blue circle), the response to the *M. leprae* sonicate challenge by group (black and orange squares) and/or the interaction analysis (blue triangle). PΔ = difference of transcript proportion between the stimulated vs non-stimulated cells by group. PΔ _T1R_ – PΔ _LEP_ = comparison of the PΔ from T1R vs LEP (interaction).

To have a better insight in DUT implicated in inflammatory responses, we selected transcripts for the genes *COMMD6*, *NKIRAS1* and *NLRP2*, which encode proteins that are critical regulators of the NF-κB pathway (Fig 4B-D). *COMMD6* encodes an inhibitor of the NF-κB pathway, modulating its transcriptional program. At baseline, LEP and T1R displayed very different *COMMD6* usage profiles (Fig 4B, S1 Table). Following *M. leprae* sonicate stimulation, we observed a strongly reduced usage of the coding transcript by the T1R group. However, the other two non-coding transcripts displayed a more dynamic usage for the LEP group (from baseline to the simulated condition, Fig 4B). This suggested that *COMMD6* was more finely regulated by LEP than T1R participants. Upon stimulation, T1R patients reached a similar profile as LEP, however given their extreme differences at baseline, T1R responded with an acute and possibly imbalanced NF-κB inhibition. This suggested transcriptome kinetics may be part of T1R pathogenesis. The *NKIRAS1* gene also encodes an inhibitor of NF-κB signaling which affects translocation of the NF-κB complex from the cytoplasm to the nucleus. *NKIRAS1* has nine annotated transcripts, however only two coding transcripts could be tested for differential usage in our data. Both transcripts encode a full-length protein with identical sequence, yet they differ in 5’ and 3’ UTR sequence and length resulting in a small size difference of 1612 bp vs 1644 bp. The two transcripts were DUT at baseline with T1R displaying a higher usage of the longer transcript. However, stimulation with *M. leprae* sonicate resulted in a highly favored usage of the shorter isoform, for both groups (Fig 4C). The preferred usage of transcripts with shorter UTRs has been identified as regulatory mechanisms, since alternative 5’ sequences can result in distinct mRNA folding while shorter 3’ regions can avoid mRNA degradation [41, 42]. Finally, *NLRP2* is a member of the NOD-like receptor (NLR) protein family. *NLRP2* expression is involved in activation of the NF-κB pathway but also regulates this pathway in a negative feedback loop [43]. Out of 19 annotated transcripts, we were able to test three for differential usage, of which two were non-coding and one was protein coding (Fig 4D). None of the transcripts differed significantly in usage at baseline between T1R and LEP participants. Following *M. leprae* sonicate stimulation, only the protein-coding transcript ENST00000448584 was significant DUT for both groups. Usage of this transcript was strongly reduced among the T1R group suggesting that *NLRP2* did not contribute to NF-kB pathway activation in this group (Fig 4D).

Contrary to the observation for DET where most transcripts responded in both groups and the differences revealed in effect sizes, the majority of interaction DUT (294/326) were group-specific (i.e. transcripts had significant usage change in response to the stimulation exclusively in the cells from one patient group) (S12 Fig). We checked the distribution of coding and non-coding transcripts for the group-specific interaction DUT (Table 4). Among transcripts with increased usage in a group, there were no significant differences for protein-coding or non-coding transcripts between the T1R and LEP groups (Table 4). Conversely, we noticed significant differences between the two groups for DUT with decreased usage after stimulation (Table 4). The T1R group displayed a higher proportion of under-used protein coding DUT while LEP had more non-coding DUT being affected by decreased usage (*P* = 2.365e-05).

**Table 4.** Distribution of transcripts by coding nature and CDS length for transcripts with group-specific usage change in response to *M. leprae* sonicate challenge.

| Transcripts | Increased usage |  | Decreased usage |  |
| --- | --- | --- | --- | --- |
|  | T1R<sup>a</sup> | LEP<sup>a</sup> | T1R | LEP |
| **TOTAL** | 69 (100%) | 56 (100%) | 95 (100%) | 74 (100%) |
| **1) Protein-coding** | **48 (70%)** | **46 (82%)** | **74 (78%)<sup>c</sup>** | **34 (46%)<sup>c</sup>** |
| Canonical | 15 (31%) | 19 (41%) | 19 (25%) | 11 (32%) |
| Non-canonical - Same length<sup>b</sup> | 6 (13%) | 5 (11%) | 16 (22%) | 2 (6%) |
| Non-canonical - Smaller<sup>b</sup> | 23 (48%) | 20 (43%) | 36 (49%) | 16 (47%) |
| Non-canonical - Longer<sup>b</sup> | 4 (8%) | 2 (5%) | 3 (4%) | 5 (15%) |
| **2) Non-coding** | **21 (30%)** | **10 (18%)** | **21 (22%)<sup>c</sup>** | **40 (54%)<sup>c</sup>** |
| lncRNA | 1 (5%) | 2 (20%) | 4 (19%) | 2 (5%) |
| Nonsense mediated decay | 5 (24%) | 2 (20%) | 6 (29%) | 4 (10%) |
| Non-stop decay | 0 | 0 | 0 | 1 (2.5%) |
| Processed transcript | 6 (28%) | 3 (30%) | 4 (19%) | 11 (28%) |
| Retained intron | 9 (43%) | 3 (30%) | 7 (33%) | 21 (52%) |
| Transcribed unprocessed pseudogene | 0 | 0 | 0 | 1 (2.5%) |

<sup>a</sup> Percentages in bold refer to the protein-coding and non-coding transcripts proportions. Remaining percentages refer to the subcategories relative to the protein-coding or non-coding separately.
<sup>b</sup> CDS length relative to the canonical transcript of the respective gene.
<sup>c</sup> Significant Fisher's test: Protein-coding vs Non-coding (DUT decreased usage in T1R & LEP), $P = 2.365e-05$ . DUT: Differentially used transcripts; LEP: leprosy T1R-free; lncRNA: long non-coding RNA; T1R: leprosy T1R-destined.

### Integration of differential transcript expression and usage in response to *M. leprae* antigen

Pooling the results from transcript expression and usage analyses revealed that roughly half of DUT overlapped with DET in the response to *M. leprae* antigens (Fig 5A). Checking which transcripts were DUT and DET for differential group response resulted in 17 transcripts (Fig 5B). From this list, we selected nine transcripts based on the functional annotation of their eight parental genes (*NLN*, *DUSP8*, *DOK1*, *OSBPL2*, *CMKLR1*, *MS4A2*, *NKRF* and *CCL5*). For instance, *NLN* is a mitochondrial peptidase that participates in clearance of leftover signal peptides that were cleaved from imported cytoplasmic proteins [44]. Disruptions in clearance processes, such as the one *NLN* is involved in, can lead to mitochondrial toxicity and cell death [44]. Following stimulation, both groups displayed increased expression of the *NLN* transcript for the canonical protein-coding and a non-coding transcript, however only the non-coding transcript displayed a significantly higher response in T1R only (Fig 5B and S2 Table). These transcripts were also DUT for both groups. The coding transcript was strongly under-used while the non-coding transcript greatly increased in usage, resulting in a usage switch event, however with twice as large changes for T1R (S5 Table). While both expression and usage of the non-coding transcript were significantly favored in T1R patients, the effect of the non-coding transcript on NLN protein expression and functional activity are unknown.

**Fig 5.**
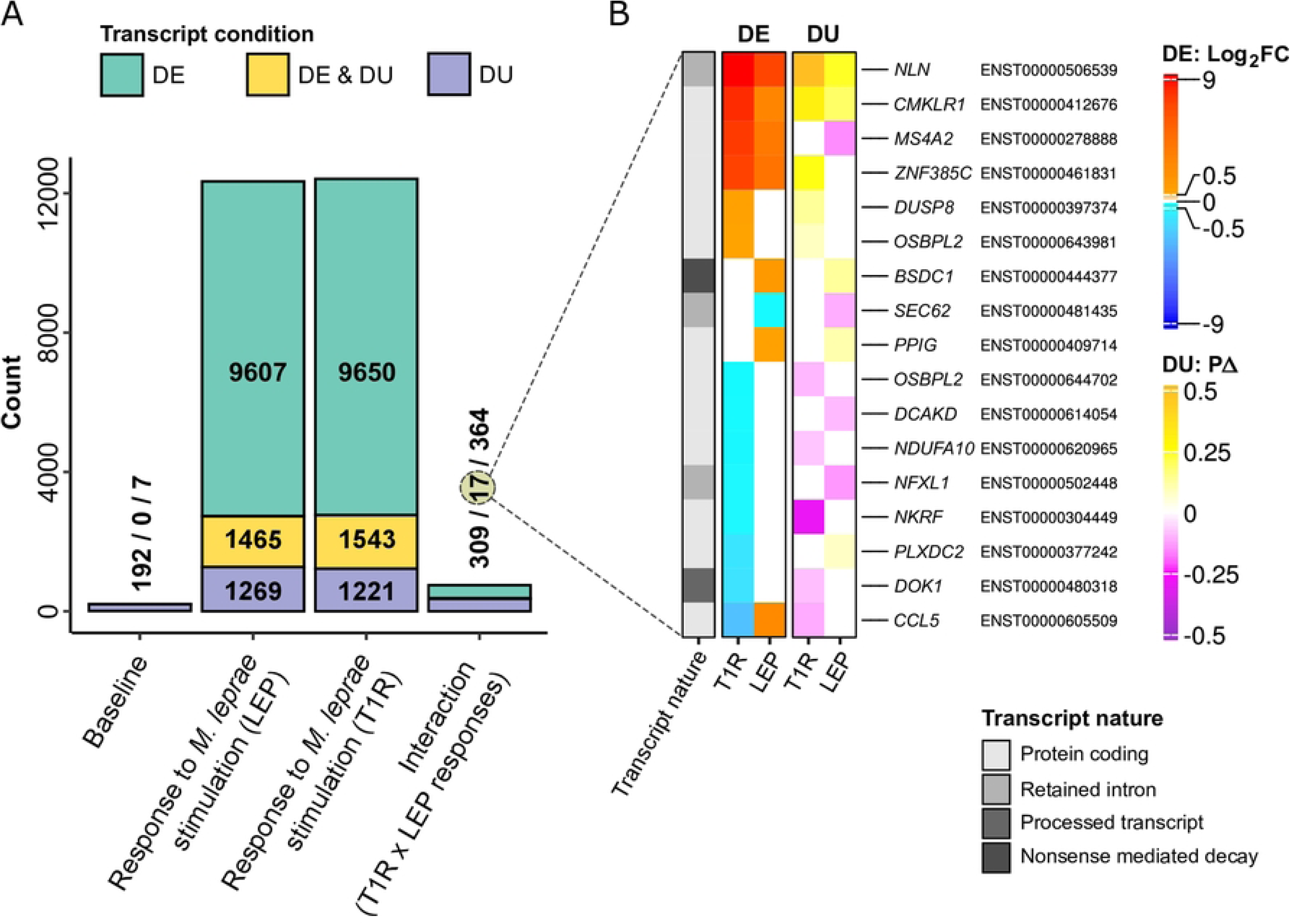
Transcripts displaying both differential expression and usage after *M. leprae* sonicate stimulation highlight the altered transcriptome of T1R-destined patients. (**A**) Barplot summarizing the number of transcripts that were differentially expressed (DE), differentially used (DU) or both for the contrasts indicated on the x-axis. (**B**) Heatmap for the mean Log_2_FC (DE) and mean PΔ (DU) for transcripts that were detected with differential expression and usage responses between groups. Shades of red and blue represent, respectively, positive and negative Log_2_FC values for individual group differential expression. Shades of yellow and magenta map the spectrum of positive to negative PΔ values for differential usage. Rows were sorted by Log_2_FC _T1R_ displayed in the first column. All 17 overlapping significant transcripts are labeled by ENSEMBL transcript ID and their parental gene symbol. The left-most column contains annotations for coding potential of each transcript or the type of non-coding transcript.

*CMKLR1* encodes a chemoattractant receptor for TIG-2 (*RARRES2*), which is expressed by M1 but not M2 macrophages and is involved in suppression of neutrophil mediated inflammation. The CMKLR1 receptor is also involved in cytokine production by macrophages via binding of its Resolvin E1 ligand [45, 46]. CMKLR1 is involved in a number of additional metabolic processes, including angiogenesis of colorectal cancers [47, 48]. Three transcripts for *CMKLR1* were detected, all significantly up-regulated with trends of stronger response in T1R. However only the protein-coding ENST00000412676 was significant for differential expression response, presenting one of the highest differences between the groups (Log_2_FC_T1R_ = 8.2 and Log_2_FC_LEP_ = 3.01, S2 Table). This transcript was, in addition, a DUT impacted by usage switch that also displayed a high usage difference in T1R patients (PΔ _T1R_ = 0.31 and PΔ _LEP_ = 0.18, S5 Table). Considering that both expression and usage were higher in T1R responses, this multipotent immune modulator is a worthwhile target for further investigation in the innate predisposition to T1R. Another immune regulatory gene, *NKRF,* encodes a transcriptional repressor of NF-κB that localizes in the nucleolus where it is a key element of nucleolar homeostasis. Disruption of nucleolar function is a common trigger for activation of the NF-κB pathway. In addition, this transcription factor directly interacts with NF-κB target genes such as *INFB*, *IL8* and *NOS2* [49, 50]. The transcript differential expression was significant only for T1R, with a moderated down-regulation (Log_2_FC_T1R_ = -0.73). However, the transcript presented a pronounced under-usage for T1R (PΔ _T1R_ = -0.29) favoring over-usage of a transcript with longer CDS, but smaller 3′ UTR region (ENST00000542113, PΔ _T1R_ = 0.29, S5 table). Since the protein coding transcript for *NKRF* is strongly suppressed in T1R patients, by expression and usage, this observation suggested the maintenance of nucleolar function as possible T1R control element. Other implicated genes such as *DUSP8*, *DOK1* and *OSBPL2* are involved in the control of signaling cascades that influence the immune response; while *MS4A2* and *CCL5* have direct effects on inflammatory processes [38, 51-54]. Why *CCL5*, which encodes a pro-inflammatory cytokine, is characterized by down-regulation and under-usage of a shorter coding transcript, while favoring usage of a transcript with much longer UTRs (ENST00000605140, Table S5) for T1R is not clear. The most parsimonious explanation may be that the opposite transcription effect of *CCL5* by the LEP group is a signal of underlying immune dysregulation in T1R patients. Some anti-inflammatory mechanisms do work, e.g. down-regulation of *CCL5*, but are overridden by other broader pro-inflammatory mechanisms (S4 Table).

## Discussion

T1R are classical T cell-mediated hypersensitivity reactions that are major contributors to nerve damage and irreversible deformities. Diagnosis of patients affected by T1R at an early stage and the initiation of interventions preventing disease progress greatly improve treatment outcomes and the quality of life for patients. Among patients with delayed diagnosis, approximately one out of four T1R patients does not respond to standard steroid therapy and severe adverse effects of extended steroid treatment often occur [55]. The need for early diagnosis of T1R patients has triggered an intense search for early onset biomarkers. While the classification of T1R severity has been improved [55], no biomarkers exist that allow the identification of leprosy patients destined for T1R before the onset of clinical symptoms.

A number of studies have probed immune mediators, both pro- and anti-inflammatory, as well as genetic polymorphisms in genes coding for those mediators or controlling their production for association with clinical T1R (rev in [4, 56]). These genetic, epidemiological, immunological, transcriptomic and metabolomic studies have provided a high-resolution view of the hypersensitive reactions underlying T1R, resulting in useful tools for its improved diagnosis. Yet, study of these immune mediators and their controlling genetic variants have not resulted in the discovery of clinically useful biomarkers for detection of T1R before the onset of clinical symptoms [56]. Similarly, expression of specific *M. leprae* genes can be used as part of the clinical diagnosis of T1R but expression levels of these pathogen genes cannot be used to identify those leprosy patients at risk of T1R [57].

A potentially more powerful approach for the identification of T1R risk biomarkers was provided by studies that followed the same patients from their leprosy diagnosis through onset of T1R and after treatment. For example, serum levels of five cytokines and monokines and three cytokine receptors were followed along T1R development but failed to identify clinically useful changes in these markers [58]. Similarly, measurements of *CXCL10* transcripts, a powerful indicator of inflammatory host responses, before, during and after T1R failed to identify predictive changes for T1R risk [59]. In a transcriptomic study, gene expression patterns of PBMC determined by RNA-seq from ten patients were followed from leprosy diagnosis, to T1R and post T1R treatment [5]. The results showed a strong impact of the host genetic background on gene expression changes revealed by strong inter-ethnicity and inter-individual differences and identified a key role for a systemic IFN-γ inducible gene signature during T1R development [5]. Unfortunately, these longitudinal changes during T1R development could not be used to derive biomarkers for T1R risk.

In a previous microarray-based transcriptomic study, we had measured the transcriptomic response to *M. leprae* sonicate of whole blood from leprosy patients at the time of leprosy diagnosis. After a follow-up of three years, a subset of those patients developed T1R and the transcriptomic response to *M. leprae* antigens of these patients was compared to that of non-T1R patients [10]. This study design had a dual purpose. It was focused on the search for biomarkers to identify those at increased risk of T1R before onset of symptoms. In addition, transcriptome response differences between the T1R and non-T1R groups at this pre-clinical stage were thought to reveal inherent mechanistic differences between the two groups in their response to *M. leprae*. Such mechanistic response differences, in turn, might highlight new targets for pharmacological interventions for prevention and/or treatment of T1R. In the present set of experiments, we maintained the intrinsic advantages of that study design but expanded on the previous approach by conducting RNA-seq and focusing the analysis on gene transcripts (i.e. isoforms).

Transcript expression and usage analyses provide different perspectives on RNA transcription dynamics that can complement each other yielding increased understanding of transcriptome changes that cannot be achieved by gene-based transcriptomics. For instance, among the transcripts significantly up-regulated by both groups in response to *M. leprae* sonicate, greater effect sizes were observed for the T1R-destined group and those transcripts clustered in terms and pathways enriched for inflammatory response and intra-cellular defense. Of note, T1R presented broad higher up-regulation of transcripts enriched for apoptotic processes, however some transcripts had higher response for LEP. Among these isoforms were *OLFM4*, *OLR1* and *PRKCD* that are apoptotic regulators. The first two genes have pro- or anti-apoptotic effects which are cell- or tissue-dependent [60-63], while *PRKCD* is pro-apoptotic [64]. Two *PRKCD* isoforms were induced by *M. leprae* sonicate stimulation, each exclusively changing in each patient group, where the transcript with shorter UTRs was the significant one for T1R. This scenario was observed for other transcripts linked to inflammatory processes, providing support for the notion that the two patient groups possess a different transcriptome response dynamic.

Additional insights were provided by the response of transcripts that implicated the genes *CASP1, CASP4, CASP5* and *CASP10*, also regulators of apoptotic pathways. For the LEP group *CASP4* was higher up-regulated and *CAPS10* was exclusively down-regulated, while T1R-destined patients presented higher up-regulation for *CASP1* and *CAPS5*. These transcripts in concert with the ones for *OLFM4*, *OLR1* and *PRKCD* suggest a balancing mechanism for LEP while T1R-destined patients presented a stronger drive towards apoptosis. The latter conclusion is consistent with the finding for the gain-of function *LRRK2* R1628P mutation which is anti-apoptotic and protective for T1R [65]. Taken together, our results suggested that disturbances in apoptosis mechanisms are a relevant factor for T1R risk, with LEP patients possessing a more controlled apoptosis program. In parallel, the most numerous transcript differences we found were related to the crosstalk between innate and adaptive immunity. For these processes, T1R clearly showed a stronger pro-inflammatory transcriptome program upon antigen challenge and our results overlapped with previous reports that investigate biopsy or whole blood of T1R patients [5, 6, 66]. Our study provided new contributions by identifying new candidate genes for T1R genesis. Of note, transcripts with varying exon, intron and UTR compositions can lead to distinct expression kinetics, and after alternative splicing, have the potential to be translated into full length proteins with alternative functions.

Our results pointed to the possible role provided by non-coding transcripts, e.g. with “retained introns.” Such transcripts may have regulatory functions for their parental gene expression [67] or even underlie pathological processes [68]. Similarly, non-coding lncRNAs have strong immune regulatory functions [69]. In our study, we observed a significantly higher up-regulation in response to *M. leprae* sonicate of transcripts retaining at least one intron and lncRNAs by the peripheral cells from T1R-destined patients compared to T1R-free patients. Given the involvement of these transcript in gene regulation, this finding is consistent with innately different immune-regulatory structures between the two groups. Two specific examples of a possible effect of non-coding transcripts on the T1R phenotype are *IL1RN* and *RELA.* Both genes participate in the control of inflammatory responses and had all of their tested transcripts up-regulated in both groups. While all transcripts displayed a stronger response by T1R patients, only their non-coding, retained-intron transcripts, were significantly different between the two groups. A key role of non-coding RNAs in T1R risk is consistent with findings of a genome-wide association analysis of T1R which identified a lncRNA gene as major risk factor [70].

In this study, it was not possible to present a transcript-specific mechanistic view of how transcripts influenced inflammatory or the innate/adaptive immunity pathways given the scarcity of knowledge about the biological function of each transcript. Nevertheless, our results presented new insight of how T1R-destined leprosy patients display a two-way impaired communication system between innate and adaptive immune responses, that ultimately leads to loop of destructive inflammatory reactions.

## Data Availability

RNA-seq raw sequences and corresponding estimated expression matrices analyzed in this work are being submitted to NCBI’s Gene Expression Omnibus repository and will be available at the time of publication.

## Acknowledgments

We are grateful to all patients who participated in the study. We thank the members of the leprosy control program at the Hospital for Dermato-Venereology in Ho Chi Minh City and the staff of the leprosy control program in Southern Việt Nam for their dedication and help in conducting the study. We thank Drs M. Tremblay, J. Pelletier (McGill University) and L.B. Barreiro (University of Chicago) for helpful comments. We also thank all members of the Schurr laboratory in Montreal and the members of the Human Genetics of Infectious Diseases laboratory in Paris for helpful discussions and suggestions.

## Supporting information

### Supplementary files

**S1 File. Supplementary methods.**

### Supplementary figures

**S1 Fig. Prospective study design and group definition.** We enrolled 32 patients at the time of leprosy diagnosis and obtained whole blood samples. Aliquots were incubated with media only (baseline, non-stimulated samples) or media + *Mycobacterium leprae* sonicate (stimulated samples), and total RNA was extracted from all samples and frozen (left box). After three years of follow-up, 22 participants remained T1R-free (LEP group) while 10 developed the T1R reactional state (T1R group). Stored RNA samples obtained at enrolment were sequenced after the three-year follow-up yielding a total of 64 RNA-seq libraries (right box).

**S2 Fig. Scheme for the *ad hoc* algorithm to identify transcript switch after Differential Transcript Usage analyses.** Boxplots in step 1 to 5 represent quartiles for the distribution of fitted usage proportion of each transcript in the corresponding RNA-seq libraries in the context of T1R-free patients. The y-axis indicates fitted proportion values while the x-axis groups the different transcripts by library type (e.g. LEP = T1R-free leprosy patients at baseline condition; LEP_st_ *M. leprae* antigen-stimulated sample). **Step 1:** For each of the library groups, we retrieved all transcripts tested for differential transcript usage, for each gene. For each group and condition separately, transcripts were sorted by their medians that were calculated from the distribution of the fitted usage proportions. **Step 2:** The fitted usage distributions of transcripts were tested via paired sample Wilcoxon tests, within each group and experimental condition. **Step 3:** Next, we assigned numeric ranks for statistically different usage distributions between two transcripts. If transcripts with different median were not significantly different, they were assigned the same ranking. **Step 4:** We retrieved the information from differential transcript usage analysis to determine which transcripts were DUT following stimulation with *M. leprae* antigen. **Step 5:** Lastly, for all transcripts, we linked the ranking information with their DUT status. If a transcript changed rank following antigen stimulation and was DUT, we called this a switch event. Finally, we checked if a DUT detected at the differential group response had different transcript switches profiles for the LEP or T1R groups following *M. leprae* antigen stimulation. If so, we also recorded a switch event.

**S3 Fig. Baseline whole blood transcriptome comparison between T1R-destined (T1R) and T1R-free leprosy patients (LEP).** (**A**) Scatterplot for differential transcript expression at baseline. All tested transcripts are represented as dots and their mean Log_2_ expression in whole blood is plotted for LEP (x-axis) vs T1R (y-axis) participants. Seven transcripts with mean expression levels that differed significantly between T1R and LEP participants are highlighted as red dots. (**B**) Transcripts with significant proportion usage difference at baseline. Results are presented as strip plot with the mean fitted proportion usage per group on the y-axis and transcripts ordered by mean proportion usage by the LEP group (shown by grey dots) on the x-axis. Blue colored dots display the mean proportion usage for the corresponding transcripts by the T1R group.

**S4 Fig. Intersection of significant terms from GO/pathway enrichment analysis for gene sets implicated by up- and down-regulated (combined) or up-regulated transcripts only.** (**A**) Venn diagram for the overlap of terms obtained by considering only up-regulated or up- and down-regulated (combined) transcripts for the group of LEP patients. (**B**) Venn diagram for overlap of terms obtained by considering only up-regulated or combined (up- and down-regulated) transcripts for the group of T1R group. (**C**) Dot plot for a selection of significant GO/pathway terms representing aspects of the immune response to *M. leprae* antigens. Dot sizes are a function of gene count (implicated by significant transcripts) and shades of gray represent negative Log_10_ (BH FDR), with darker colors meaning higher significance.

**S5 Fig. Manhattan plots for the results of pathway and GO-term enrichment tests on differentially expressed transcript (DET) subsets by effect size differences between T1R and LEP responses.** (**A**) Manhattan plot for GO-terms, KEGG and Reactome pathways with significant enrichment of the 2948 DET with intermediate effect size difference between T1R and LEP responses (0.2 > |Log_2_FC| < 0.5; Fig 2 C, yellow dots). (**B**) Manhattan plot for GO-terms, KEGG and Reactome pathways with significant enrichment of the 1190 DET with |Log_2_FC| ≥ 0.5 response difference between the T1R and LEP groups (Fig 2 C, green dots). For A and B, the y-axis indicates the negative Log_10_ for BH adjusted P-values for the terms arranged along the x-axis. The horizontal dashed line represents the 10% FDR cut-off for significant pathways/GO-terms. Dots are sized as a function of the gene number in a term and colors represent the database from which the terms were obtained.

**S6 Fig. Group-specific up-regulated transcripts show overlap in biological functions, but also reveal group differences in the response to *M. leprae* antigens.** (**A**) Venn diagram for the number of significantly enriched GO terms and pathways identified by testing lists of genes implicated by positive Log_2_FC of differentially expressed transcripts (DET) exclusive to T1R (465) or LEP (574). (**B**) Barplot for a selection of significant terms that represent intracellular defense/immune response among the107 overlapping terms from panel A. The left panel shows results for the LEP group, while the right panel refers to T1R. Terms were arranged along the y-axis as function of increasing Benjamini-Hochberg FDR for the LEP group. The x-axis displays the gene count for each term. (**C**) Barplots for LEP-specific terms (left panel) and T1R-specific ones (right). Terms sorting was done for each group by increasing Benjamini-Hochberg FDR values. Highlighted in yellow is an example of a physiologically similar term that was found among shared and group specific terms indicating a common biological theme across group-specific responses. The grey-shaded term represents an example of a LEP-specific term which was not enriched in T1R, while green-shaded refers to T1R-specific enrichment.

**S7 Fig. Transcript response differences between T1R and LEP patients for GO terms, KEGG or Reactome pathways that showed significant enrichment for both the LEP and T1R groups.** Immune response-linked terms for which the 5639 overlapping up-regulated DET presented in Figure 2D and 2E were significantly enriched. **(A)** Dotplots of group mean Log_2_FC of all DET of a given term. (**B**) Dotplots of the sum of transcript-wise residual Log_2_FC difference for the LEP and T1R groups. This metric was derived as single value per term and per group as followed: i) Identify the group with the higher Log_2_FC for a given transcript; ii) the group with the higher value is assigned the difference Log_2_FC _A_ - Log_2_FC _B_ while the other group is assigned zero for this transcript. The result for each term is a two-column matrix, each column representing a group, with as many rows as DET in a term enriched; iii) for each group, summing the column of transcript values resulted in one value per term and group. (**C**) Heatmap for all DET of the GO term “inflammatory response”. The mean Log_2_FC for all DET was retrieved for each group and ordered by decreasing values of the LEP group. This example shows how the residual Log_2_FC for enriched terms more strongly displays subtle differences in transcriptional response of the two groups.

**S8 Fig. Profiles for transcripts with significant differential response between the T1R and LEP groups after stimulation with *M. leprae* sonicate.** (**A)** Classes of differentially expressed transcripts (DET) in response to *M. leprae* sonicate detected via interaction analysis. The effect size difference between two groups is commonly expressed as +/- Log_2_FC as shown in the examples column. Response differences were grouped into six categories as illustrated here. Transcript details are shown in S3 table. (**B-F**) Strip plots presenting the Log_2_FC_T1R_ and Log_2_FC_LEP_ from 381 significant differentially expressed transcripts (Fig 3A), separated according to the direction of the changes: **B**) up-regulated in the T1R and down-regulated in the LEP group (black and green dots), **C**) down-regulated in both groups but more in the LEP group (black and green dots), **D**) down-regulated in both groups but more in the T1R group (black and purple dots), **E**) down-regulated in the T1R group and up-regulated in the LEP group (black and purple dots) and **F**) up-regulated in both groups but more in the in LEP group (black and purple dots). On the x-axis, transcripts are ordered by their Log_2_FC_LEP_. Effect size is represented as mean Log_2_FC per group (y-axis). Grey dots represent Log_2_FC_LEP_. Colored dots display the Log2FC_T1R._ Open dots are Log_2_FC for both groups that do not fall in the category presented. The number of transcripts in each category is shown in bold on top of each panel. For panels B, C, and E the majority of corresponding gene symbols are shown for DET. For panels D and F, labeled dots identify transcripts of genes encoding proteins with immunologic/inflammatory functions.

**S9 Fig. Stimulation with *M. leprae* sonicate induces higher activation of transcripts related to intracellular response and inflammatory processes in T1R-destined leprosy patients versus T1R-free leprosy patients.** Heatmap for 145 transcripts detected with significant differential response to *M. leprae* antigens between T1R and LEP patients. Columns present (**A**) all 145 up-regulated DET with stronger response by T1R, (**B**) transcripts representing the inflammatory response, (**C**) transcripts that represent the response to interferon-gamma, and (**D**) transcripts that represent the NOD-like receptor signaling pathway. The main columns A to D are composed of two sub-columns representing the responses for the T1R or LEP groups. Rows represent the same transcript across all columns and are assigned to group 1 if their parental gene is protein coding or to group 2 if transcripts were annotated to non-coding genes. Colors represent Log_2_FC values, with strength of responses depicted by scales of red. The right most column present annotations for transcript nature.

**S10 Fig. Effect size distribution and intersections of transcript usage response upon stimulation with *M. leprae* sonicate by T1R-free (LEP) and T1R-destined (T1R) leprosy patients.** (**A**) Strip plot presenting the distribution of the mean fitted proportion difference (PΔ) between stimulated and non-stimulated samples by group. Transcripts represented by dots were ordered along the x-axis according to increasing PΔ values for the LEP group. Light gray dots represent transcripts with non-significant PΔ for both groups and horizontal lines mark the | PΔ | ≥ 0.07 threshold. Differentially used transcripts (DUT) for LEP are depicted as black dots and appear as a continuous curve in the center part of the graph while colored dots display PΔ for the T1R group. Red dots show transcripts for which the response difference between groups (interaction) was below the 0.07 cut-off (i.e. |PΔ_T1R_ – PΔ_LEP_|< 0.07), yellow represent DUT with 0.07 ≤ |PΔ| < 0.1 and green dots for |PΔ| ≥ 0.1. Colored numbers represent the counts of DUT for the corresponding PΔ intervals. Yellow and green dots are significant interaction DUT, with the green dots indicating the top 30% of the percentile distribution of significant DUT. (**B**) UpSet plot presenting the differential transcript usage for LEP and T1R responses. The bar chart on the bottom left indicates the total number of increased (over) or decreased (under) transcript usage by the two groups, while the bottom panel identifies how sets intersected. The vertical upper bars indicate the corresponding intersection size (transcript counts). Connected dots indicate DUT that overlap between the categories (sets) shown on the left while unconnected dots highlight DUT detected only for the indicated group.

**S11 Fig. Heatmap for 150 differentially used transcripts (DUT) annotated to immune or inflammatory functions with highest response difference between LEP and T1R.** We i) retrieved 1281 DUT that belonged to genes with annotated immunological functions, ii) derived the difference of group responses (i.e. PΔ_T1R_ – PΔ_LEP_), iii) sorted the absolute values and selected the top 150 DUT with the largest differences. Next, we selected three GO terms to present how DUT effect sizes compared between groups and how DUT overlapped in the selected terms. For the three main columns, each contains two sub-columns representing effect sizes for the T1R or LEP groups, where PΔ of DUT are listed. Column (**A)** presents DUT that were annotated to the GO term “inflammatory response”, (**B)** represents the “innate immune response” GO term and (**C)** displays DUT for the GO term “apoptotic process”. Each row corresponds to the same DUT across the entire plot since the same gene/transcript may be part of more than one term. Colored cells highlight significant PΔ for a transcript. Non-significant transcripts or ones that were not part of the three selected terms had their PΔ values set to zero (white colored). Shades of red represent positive PΔ values (over-usage in relation to the baseline for each group) and shades of blue represent negative PΔ (under-usage). Transcript biotypes are indicated by a color scheme listed on the right. Gene symbols are shown for each transcript.

**S12 Fig. Differentially used transcripts (DUT) for T1R and LEP groups suggest dysregulation of inflammatory circuitry in T1R.** This plot presents the usage response profiles for the 326 differentially used transcripts detected via interaction analysis. The two central columns display the PΔ for the T1R and LEP groups after stimulation with *M. leprae* sonicate. Shades of red represent positive PΔ values (over-usage) and shades of blue represent negative PΔ (under-usage). Group-specific interaction DUTs (n = 294) responded to the stimulation with differential usage exclusively in one group. They are shown in white for the group where the response was not significant and colored for the group where the response was significant. The remaining interaction DUTs responded to the stimulation in both groups in the same direction but with significantly different size effect (n = 28) or in opposite direction (n =4), as depicted by the different color shades in the two columns. Rows were sorted by decreasing values of PΔ_T1R_, then re-arranged in three blocks: i) positive PΔ for both groups, ii) opposite PΔ and iii) negative PΔ for both groups. Next, DUT with non-significant changes were assigned PΔ = 0. Given that no gene ontology (GO) or pathway enrichments were found, we searched the annotated function of each implicated gene in the GO, Reactome and KEGG databases. In this process, we detected five major annotation themes: immune response, inflammatory processes, apoptosis, ubiquitination and RNA processing. Transcripts that represented any of these categories are labeled by their gene symbol and the GO/pathway tagged by those genes (colored circles). The left-most column contains annotations for each transcripts’ coding potential or type of non-coding transcript. Gene symbols in red case present examples discussed in the main text, while bold letters indicate transcripts displaying a usage switch. PΔ = difference of transcript proportion between the stimulated vs non-stimulated cells by group.

### Supplementary tables

**S1 Table.** Differential transcript usage results for baseline differences between T1R against LEP.

**S2 Table.** Differential transcript expression results for all contrasts addressing stimulation effect.

**S3 Table.** Profiles for transcripts with significant *M. leprae* response difference between the T1R and LEP groups.

**S4 Table.** Results for gene ontology and pathway enrichment analysis for 145 transcripts with higher up-regulation in T1R group.

**S5 Table.** Differential transcript usage results for all contrasts addressing stimulation effect.

### Supplementary data

**S1 Data.** Intersection of significant Gene Ontology, Reactome and KEGG terms resulting from testing all DET for LEP or T1R responses to *M. leprae* stimulation.

## References

1. Scollard DM, Adams LB, Gillis TP, Krahenbuhl JL, Truman RW, Williams DL. The continuing challenges of leprosy. Clin Microbiol Rev. 2006;19(2):338–81. Epub 2006/04/15. doi: 10.1128/cmr.19.2.338-381.2006. PubMed PMID: 16614253; PubMed Central PMCID: PMCPMC1471987.

2. World Health Organization = Organisation mondiale de la S. Global leprosy (Hansen disease) update, 2020: impact of COVID-19 on global leprosy control –Situation de la lèpre (maladie de Hansen) dans le monde, 2020: impact de la COVID-19 sur les activités mondiales de lutte contre la lèpre. Weekly Epidemiological Record = Relevé épidémiologique hebdomadaire. 2021;96(36):421-44.

3. Fava V, Orlova M, Cobat A, Alcaïs A, Mira M, Schurr E. Genetics of leprosy reactions: an overview. Mem Inst Oswaldo Cruz. 2012;107 Suppl 1:132–42. Epub 2013/01/11. doi: 10.1590/s0074-02762012000900020. PubMed PMID: 23283464.

4. Geluk A. Correlates of immune exacerbations in leprosy. Semin Immunol. 2018;39:111–8. Epub 2018/06/29. doi: 10.1016/j.smim.2018.06.003. PubMed PMID: 29950273.

5. Teles RMB, Lu J, Tió-Coma M, Goulart IMB, Banu S, Hagge D, et al. Identification of a systemic interferon-γ inducible antimicrobial gene signature in leprosy patients undergoing reversal reaction. PLoS Negl Trop Dis. 2019;13(10):e0007764. Epub 2019/10/11. doi: 10.1371/journal.pntd.0007764. PubMed PMID: 31600201; PubMed Central PMCID: PMCPMC6805014.

6. Tió-Coma M, van Hooij A, Bobosha K, van der Ploeg-van Schip JJ, Banu S, Khadge S, et al. Whole blood RNA signatures in leprosy patients identify reversal reactions before clinical onset: a prospective, multicenter study. Sci Rep. 2019;9(1):17931. Epub 2019/12/01. doi: 10.1038/s41598-019-54213-y. PubMed PMID: 31784594; PubMed Central PMCID: PMCPMC6884598.

7. World Health O. Towards zero leprosy. Global leprosy (Hansen’s Disease) strategy 2021–2030. New Delhi: World Health Organization; 2021 2021.

8. Fava VM, Dallmann-Sauer M, Schurr E. Genetics of leprosy: today and beyond. Hum Genet. 2020;139(6-7):835–46. Epub 2019/11/13. doi: 10.1007/s00439-019-02087-5. PubMed PMID: 31713021.

9. Freuer D, Meisinger C. Association between inflammatory bowel disease and Parkinson’s disease: A Mendelian randomization study. NPJ Parkinsons Dis. 2022;8(1):55. Epub 2022/05/10. doi: 10.1038/s41531-022-00318-7. PubMed PMID: 35534507; PubMed Central PMCID: PMCPMC9085764.

10. Orlova M, Cobat A, Huong NT, Ba NN, Van Thuc N, Spencer J, et al. Gene set signature of reversal reaction type I in leprosy patients. PLoS Genet. 2013;9(7):e1003624. Epub 2013/07/23. doi: 10.1371/journal.pgen.1003624. PubMed PMID: 23874223; PubMed Central PMCID: PMCPMC3708838.

11. Schroeder A, Mueller O, Stocker S, Salowsky R, Leiber M, Gassmann M, et al. The RIN: an RNA integrity number for assigning integrity values to RNA measurements. BMC Mol Biol. 2006;7:3. Epub 2006/02/02. doi: 10.1186/1471-2199-7-3. PubMed PMID: 16448564; PubMed Central PMCID: PMCPMC1413964.

12. Manry J, Nédélec Y, Fava VM, Cobat A, Orlova M, Thuc NV, et al. Deciphering the genetic control of gene expression following Mycobacterium leprae antigen stimulation. PLoS Genet. 2017;13(8):e1006952. Epub 2017/08/10. doi: 10.1371/journal.pgen.1006952. PubMed PMID: 28793313; PubMed Central PMCID: PMCPMC5565194.

13. Andrews S. FastQC: A Quality Control Tool for High Throughput Sequence Data [Online]. Available online at: http://www.bioinformatics.babraham.ac.uk/projects/fastqc/. 2010.

14. Wang L, Wang S, Li W. RSeQC: quality control of RNA-seq experiments. Bioinformatics. 2012;28(16):2184–5. Epub 2012/06/30. doi: 10.1093/bioinformatics/bts356. PubMed PMID: 22743226.

15. Martin M. Cutadapt Removes Adapter Sequences From High-Throughput Sequencing Reads. EMBnetjournal. 2011. doi: 10.14806/ej.17.1.200.

16. Dobin A, Davis CA, Schlesinger F, Drenkow J, Zaleski C, Jha S, et al. STAR: ultrafast universal RNA-seq aligner. Bioinformatics. 2013;29(1):15–21. Epub 2012/10/30. doi: 10.1093/bioinformatics/bts635. PubMed PMID: 23104886; PubMed Central PMCID: PMCPMC3530905.

17. Cunningham F, Allen JE, Allen J, Alvarez-Jarreta J, Amode MR, Armean IM, et al. Ensembl 2022. Nucleic acids research. 2022;50(D1):D988-d95. Epub 2021/11/19. doi: 10.1093/nar/gkab1049. PubMed PMID: 34791404; PubMed Central PMCID: PMCPMC8728283.

18. Patro R, Duggal G, Love MI, Irizarry RA, Kingsford C. Salmon provides fast and bias-aware quantification of transcript expression. Nat Methods. 2017;14(4):417–9. Epub 2017/03/07. doi: 10.1038/nmeth.4197. PubMed PMID: 28263959; PubMed Central PMCID: PMCPMC5600148.

19. (2021) RCT. R: A language and environment for statistical computing. Vienna, Austria: R Foundation for Statistical Computing; 2021.

20. Soneson C, Love MI, Robinson MD. Differential analyses for RNA-seq: transcript-level estimates improve gene-level inferences. F1000Research. 2015;4:1521. Epub 2016/03/01. doi: 10.12688/f1000research.7563.2. PubMed PMID: 26925227; PubMed Central PMCID: PMCPMC4712774.

21. Durinck S, Spellman PT, Birney E, Huber W. Mapping identifiers for the integration of genomic datasets with the R/Bioconductor package biomaRt. Nature protocols. 2009;4(8):1184-91. Epub 2009/07/21. doi: 10.1038/nprot.2009.97. PubMed PMID: 19617889; PubMed Central PMCID: PMCPMC3159387.

22. Love MI, Soneson C, Patro R. Swimming downstream: statistical analysis of differential transcript usage following Salmon quantification. F1000Research. 2018;7:952. Epub 2018/10/30. doi: 10.12688/f1000research.15398.3. PubMed PMID: 30356428; PubMed Central PMCID: PMCPMC6178912.3.

23. Robinson MD, McCarthy DJ, Smyth GK. edgeR: a Bioconductor package for differential expression analysis of digital gene expression data. Bioinformatics. 2010;26(1):139–40. Epub 2009/11/17. doi: 10.1093/bioinformatics/btp616. PubMed PMID: 19910308; PubMed Central PMCID: PMCPMC2796818.

24. Robinson MD, Oshlack A. A scaling normalization method for differential expression analysis of RNA-seq data. Genome Biol. 2010;11(3):R25. Epub 2010/03/04. doi: 10.1186/gb-2010-11-3-r25. PubMed PMID: 20196867; PubMed Central PMCID: PMCPMC2864565.

25. Ritchie ME, Phipson B, Wu D, Hu Y, Law CW, Shi W, et al. limma powers differential expression analyses for RNA-sequencing and microarray studies. Nucleic acids research. 2015;43(7):e47. Epub 2015/01/22. doi: 10.1093/nar/gkv007. PubMed PMID: 25605792; PubMed Central PMCID: PMCPMC4402510.

26. Law CW, Chen Y, Shi W, Smyth GK. voom: Precision weights unlock linear model analysis tools for RNA-seq read counts. Genome Biol. 2014;15(2):R29. Epub 2014/02/04. doi: 10.1186/gb-2014-15-2-r29. PubMed PMID: 24485249; PubMed Central PMCID: PMCPMC4053721.

27. Law CW, Alhamdoosh M, Su S, Dong X, Tian L, Smyth GK, et al. RNA-seq analysis is easy as 1-2-3 with limma, Glimma and edgeR. F1000Research. 2016;5. Epub 2016/06/17. doi: 10.12688/f1000research.9005.3. PubMed PMID: 27441086; PubMed Central PMCID: PMCPMC4937821.

28. Van den Berge K, Soneson C, Robinson MD, Clement L. stageR: a general stage-wise method for controlling the gene-level false discovery rate in differential expression and differential transcript usage. Genome Biol. 2017;18(1):151. Epub 2017/08/09. doi: 10.1186/s13059-017-1277-0. PubMed PMID: 28784146; PubMed Central PMCID: PMCPMC5547545.

29. Nowicka M, Robinson MD. DRIMSeq: a Dirichlet-multinomial framework for multivariate count outcomes in genomics. F1000Research. 2016;5:1356. Epub 2017/01/24. doi: 10.12688/f1000research.8900.2. PubMed PMID: 28105305; PubMed Central PMCID: PMCPMC5200948.

30. Yu G, Wang LG, Han Y, He QY. clusterProfiler: an R package for comparing biological themes among gene clusters. OMICS. 2012;16(5):284–7. Epub 2012/03/30. doi: 10.1089/omi.2011.0118. PubMed PMID: 22455463; PubMed Central PMCID: PMCPMC3339379.

31. Yu G, He QY. ReactomePA: an R/Bioconductor package for reactome pathway analysis and visualization. Mol Biosyst. 2016;12(2):477–9. Epub 2015/12/15. doi: 10.1039/c5mb00663e. PubMed PMID: 26661513.

32. Kanehisa M, Goto S, Sato Y, Furumichi M, Tanabe M. KEGG for integration and interpretation of large-scale molecular data sets. Nucleic acids research. 2012;40(Database issue):D109-14. Epub 2011/11/15. doi: 10.1093/nar/gkr988. PubMed PMID: 22080510; PubMed Central PMCID: PMCPMC3245020.

33. Gillespie M, Jassal B, Stephan R, Milacic M, Rothfels K, Senff-Ribeiro A, et al. The reactome pathway knowledgebase 2022. Nucleic acids research. 2022;50(D1):D687-d92. Epub 2021/11/18. doi: 10.1093/nar/gkab1028. PubMed PMID: 34788843; PubMed Central PMCID: PMCPMC8689983.

34. Ashburner M, Ball CA, Blake JA, Botstein D, Butler H, Cherry JM, et al. Gene ontology: tool for the unification of biology. The Gene Ontology Consortium. Nat Genet. 2000;25(1):25–9. Epub 2000/05/10. doi: 10.1038/75556. PubMed PMID: 10802651; PubMed Central PMCID: PMCPMC3037419.

35. The Gene Ontology resource: enriching a GOld mine. Nucleic acids research. 2021;49(D1):D325-d34. Epub 2020/12/09. doi: 10.1093/nar/gkaa1113. PubMed PMID: 33290552; PubMed Central PMCID: PMCPMC7779012.

36. Ranque B, Nguyen VT, Vu HT, Nguyen TH, Nguyen NB, Pham XK, et al. Age is an important risk factor for onset and sequelae of reversal reactions in Vietnamese patients with leprosy. Clin Infect Dis. 2007;44(1):33–40. Epub 2006/12/05. doi: 10.1086/509923. PubMed PMID: 17143812.

37. Curtin JF, Cotter TG. JNK regulates HIPK3 expression and promotes resistance to Fas-mediated apoptosis in DU 145 prostate carcinoma cells. J Biol Chem. 2004;279(17):17090–100. Epub 2004/02/10. doi: 10.1074/jbc.M307629200. PubMed PMID: 14766760.

38. Amo G, García-Menaya J, Campo P, Cordobés C, Plaza Serón MC, Ayuso P, et al. A Nonsynonymous FCER1B SNP is Associated with Risk of Developing Allergic Rhinitis and with IgE Levels. Sci Rep. 2016;6:19724. Epub 2016/01/23. doi: 10.1038/srep19724. PubMed PMID: 26792385; PubMed Central PMCID: PMCPMC4726269.

39. Jacob AG, Smith CWJ. Intron retention as a component of regulated gene expression programs. Hum Genet. 2017;136(9):1043–57. Epub 2017/04/10. doi: 10.1007/s00439-017-1791-x. PubMed PMID: 28391524; PubMed Central PMCID: PMCPMC5602073.

40. Statello L, Guo CJ, Chen LL, Huarte M. Gene regulation by long non-coding RNAs and its biological functions. Nat Rev Mol Cell Biol. 2021;22(2):96–118. Epub 2020/12/24. doi: 10.1038/s41580-020-00315-9. PubMed PMID: 33353982; PubMed Central PMCID: PMCPMC7754182.

41. Leppek K, Das R, Barna M. Functional 5’ UTR mRNA structures in eukaryotic translation regulation and how to find them. Nat Rev Mol Cell Biol. 2018;19(3):158–74. Epub 2017/11/23. doi: 10.1038/nrm.2017.103. PubMed PMID: 29165424; PubMed Central PMCID: PMCPMC5820134.

42. Pai AA, Baharian G, Pagé Sabourin A, Brinkworth JF, Nédélec Y, Foley JW, et al. Widespread Shortening of 3’ Untranslated Regions and Increased Exon Inclusion Are Evolutionarily Conserved Features of Innate Immune Responses to Infection. PLoS Genet. 2016;12(9):e1006338. Epub 2016/10/01. doi: 10.1371/journal.pgen.1006338. PubMed PMID: 27690314; PubMed Central PMCID: PMCPMC5045211.

43. Rossi MN, Pascarella A, Licursi V, Caiello I, Taranta A, Rega LR, et al. NLRP2 Regulates Proinflammatory and Antiapoptotic Responses in Proximal Tubular Epithelial Cells. Front Cell Dev Biol. 2019;7:252. Epub 2019/11/12. doi: 10.3389/fcell.2019.00252. PubMed PMID: 31709256; PubMed Central PMCID: PMCPMC6822264.

44. Teixeira PF, Masuyer G, Pinho CM, Branca RMM, Kmiec B, Wallin C, et al. Mechanism of Peptide Binding and Cleavage by the Human Mitochondrial Peptidase Neurolysin. J Mol Biol. 2018;430(3):348–62. Epub 2017/12/01. doi: 10.1016/j.jmb.2017.11.011. PubMed PMID: 29183787.

45. Cash JL, Bena S, Headland SE, McArthur S, Brancaleone V, Perretti M. Chemerin15 inhibits neutrophil-mediated vascular inflammation and myocardial ischemia-reperfusion injury through ChemR23. EMBO Rep. 2013;14(11):999–1007. Epub 2013/09/04. doi: 10.1038/embor.2013.138. PubMed PMID: 23999103; PubMed Central PMCID: PMCPMC3818079.

46. Herová M, Schmid M, Gemperle C, Hersberger M. ChemR23, the receptor for chemerin and resolvin E1, is expressed and functional on M1 but not on M2 macrophages. J Immunol. 2015;194(5):2330–7. Epub 2015/02/01. doi: 10.4049/jimmunol.1402166. PubMed PMID: 25637017.

47. Wittamer V, Franssen JD, Vulcano M, Mirjolet JF, Le Poul E, Migeotte I, et al. Specific recruitment of antigen-presenting cells by chemerin, a novel processed ligand from human inflammatory fluids. J Exp Med. 2003;198(7):977–85. Epub 2003/10/08. doi: 10.1084/jem.20030382. PubMed PMID: 14530373; PubMed Central PMCID: PMCPMC2194212.

48. Kiczmer P, Seńkowska AP, Kula A, Dawidowicz M, Strzelczyk JK, Zajdel EN, et al. Assessment of CMKLR1 level in colorectal cancer and its correlation with angiogenic markers. Exp Mol Pathol. 2020;113:104377. Epub 2020/01/14. doi: 10.1016/j.yexmp.2020.104377. PubMed PMID: 31926977.

49. Coccia M, Rossi A, Riccio A, Trotta E, Santoro MG. Human NF-κB repressing factor acts as a stress-regulated switch for ribosomal RNA processing and nucleolar homeostasis surveillance. Proc Natl Acad Sci U S A. 2017;114(5):1045–50. Epub 2017/01/18. doi: 10.1073/pnas.1616112114. PubMed PMID: 28096332; PubMed Central PMCID: PMCPMC5293105.

50. Nourbakhsh M, Hauser H. Constitutive silencing of IFN-beta promoter is mediated by NRF (NF-kappaB-repressing factor), a nuclear inhibitor of NF-kappaB. Embo j. 1999;18(22):6415–25. Epub 1999/11/24. doi: 10.1093/emboj/18.22.6415. PubMed PMID: 10562553; PubMed Central PMCID: PMCPMC1171704.

51. Ding T, Zhou Y, Long R, Chen C, Zhao J, Cui P, et al. DUSP8 phosphatase: structure, functions, expression regulation and the role in human diseases. Cell Biosci. 2019;9:70. Epub 2019/08/31. doi: 10.1186/s13578-019-0329-4. PubMed PMID: 31467668; PubMed Central PMCID: PMCPMC6712826.

52. Downer EJ, Johnston DG, Lynch MA. Differential role of Dok1 and Dok2 in TLR2-induced inflammatory signaling in glia. Mol Cell Neurosci. 2013;56:148–58. Epub 2013/05/11. doi: 10.1016/j.mcn.2013.04.007. PubMed PMID: 23659921.

53. Koponen A, Pan G, Kivelä AM, Ralko A, Taskinen JH, Arora A, et al. ORP2, a cholesterol transporter, regulates angiogenic signaling in endothelial cells. Faseb j. 2020;34(11):14671–94. Epub 2020/09/12. doi: 10.1096/fj.202000202R. PubMed PMID: 32914503.

54. Zeng Z, Lan T, Wei Y, Wei X. CCL5/CCR5 axis in human diseases and related treatments. Genes Dis. 2022;9(1):12–27. Epub 2021/09/14. doi: 10.1016/j.gendis.2021.08.004. PubMed PMID: 34514075; PubMed Central PMCID: PMCPMC8423937.

55. Lockwood DN, Darlong J, Govindharaj P, Kurian R, Sundarrao P, John AS. AZALEP a randomized controlled trial of azathioprine to treat leprosy nerve damage and Type 1 reactions in India: Main findings. PLoS Negl Trop Dis. 2017;11(3):e0005348. Epub 2017/03/31. doi: 10.1371/journal.pntd.0005348. PubMed PMID: 28358815; PubMed Central PMCID: PMCPMC5373510.

56. Luo Y, Kiriya M, Tanigawa K, Kawashima A, Nakamura Y, Ishii N, et al. Host-Related Laboratory Parameters for Leprosy Reactions. Front Med (Lausanne). 2021;8:694376. Epub 2021/11/09. doi: 10.3389/fmed.2021.694376. PubMed PMID: 34746168; PubMed Central PMCID: PMCPMC8568883.

57. Sharma R, Lavania M, Chauhan DS, Katoch K, Amresh, Pramod, et al. Potential of a metabolic gene (accA3) of M. leprae as a marker for leprosy reactions. Indian J Lepr. 2009;81(3):141–8. Epub 2010/06/01. PubMed PMID: 20509343.

58. Faber WR, Iyer AM, Fajardo TT, Dekker T, Villahermosa LG, Abalos RM, et al. Serial measurement of serum cytokines, cytokine receptors and neopterin in leprosy patients with reversal reactions. Lepr Rev. 2004;75(3):274–81. Epub 2004/10/29. PubMed PMID: 15508904.

59. Scollard DM, Chaduvula MV, Martinez A, Fowlkes N, Nath I, Stryjewska BM, et al. Increased CXC ligand 10 levels and gene expression in type 1 leprosy reactions. Clin Vaccine Immunol. 2011;18(6):947–53. Epub 2011/04/22. doi: 10.1128/cvi.00042-11. PubMed PMID: 21508169; PubMed Central PMCID: PMCPMC3122607.

60. Liu RH, Yang MH, Xiang H, Bao LM, Yang HA, Yue LW, et al. Depletion of OLFM4 gene inhibits cell growth and increases sensitization to hydrogen peroxide and tumor necrosis factor-alpha induced-apoptosis in gastric cancer cells. J Biomed Sci. 2012;19(1):38. Epub 2012/04/05. doi: 10.1186/1423-0127-19-38. PubMed PMID: 22471589; PubMed Central PMCID: PMCPMC3359197.

61. Liu W, Liu Y, Li H, Rodgers GP. Olfactomedin 4 contributes to hydrogen peroxide-induced NADPH oxidase activation and apoptosis in mouse neutrophils. Am J Physiol Cell Physiol. 2018;315(4):C494-c501. Epub 2018/06/28. doi: 10.1152/ajpcell.00336.2017. PubMed PMID: 29949402; PubMed Central PMCID: PMCPMC6230683.

62. Li C, He K, Yin M, Zhang Q, Lin J, Niu Y, et al. LOX-1 Regulates Neutrophil Apoptosis and Fungal Load in A. Fumigatus Keratitis. Curr Eye Res. 2021;46(12):1800–11. Epub 2021/07/16. doi: 10.1080/02713683.2021.1948063. PubMed PMID: 34264144.

63. Khaidakov M, Mitra S, Kang BY, Wang X, Kadlubar S, Novelli G, et al. Oxidized LDL receptor 1 (OLR1) as a possible link between obesity, dyslipidemia and cancer. PLoS One. 2011;6(5):e20277. Epub 2011/06/04. doi: 10.1371/journal.pone.0020277. PubMed PMID: 21637860; PubMed Central PMCID: PMCPMC3102697.

64. Baumann U, Fernández-Sáiz V, Rudelius M, Lemeer S, Rad R, Knorn AM, et al. Disruption of the PRKCD-FBXO25-HAX-1 axis attenuates the apoptotic response and drives lymphomagenesis. Nat Med. 2014;20(12):1401–9. Epub 2014/11/25. doi: 10.1038/nm.3740. PubMed PMID: 25419709.

65. Fava VM, Xu YZ, Lettre G, Van Thuc N, Orlova M, Thai VH, et al. Pleiotropic effects for Parkin and LRRK2 in leprosy type-1 reactions and Parkinson’s disease. Proc Natl Acad Sci U S A. 2019;116(31):15616–24. Epub 2019/07/17. doi: 10.1073/pnas.1901805116. PubMed PMID: 31308240; PubMed Central PMCID: PMCPMC6681704.

66. Teles RM, Graeber TG, Krutzik SR, Montoya D, Schenk M, Lee DJ, et al. Type I interferon suppresses type II interferon-triggered human anti-mycobacterial responses. Science. 2013;339(6126):1448-53. Epub 2013/03/02. doi: 10.1126/science.1233665. PubMed PMID: 23449998; PubMed Central PMCID: PMCPMC3653587.

67. Schmitz U, Pinello N, Jia F, Alasmari S, Ritchie W, Keightley MC, et al. Intron retention enhances gene regulatory complexity in vertebrates. Genome Biol. 2017;18(1):216. Epub 2017/11/17. doi: 10.1186/s13059-017-1339-3. PubMed PMID: 29141666; PubMed Central PMCID: PMCPMC5688624.

68. Zhang J, Liu H, Liu Z, Liao Y, Guo L, Wang H, et al. A functional alternative splicing mutation in AIRE gene causes autoimmune polyendocrine syndrome type 1. PLoS One. 2013;8(1):e53981. Epub 2013/01/24. doi: 10.1371/journal.pone.0053981. PubMed PMID: 23342054; PubMed Central PMCID: PMCPMC3540864.

69. Chen YG, Satpathy AT, Chang HY. Gene regulation in the immune system by long noncoding RNAs. Nat Immunol. 2017;18(9):962–72. Epub 2017/08/23. doi: 10.1038/ni.3771. PubMed PMID: 28829444; PubMed Central PMCID: PMCPMC9830650.

70. Fava VM, Manry J, Cobat A, Orlova M, Van Thuc N, Moraes MO, et al. A genome wide association study identifies a lncRna as risk factor for pathological inflammatory responses in leprosy. PLoS Genet. 2017;13(2):e1006637. Epub 2017/02/22. doi: 10.1371/journal.pgen.1006637. PubMed PMID: 28222097; PubMed Central PMCID: PMCPMC5340414.

